# Drivers of Oncologist Preference of AI-Generated Literature Review in a Randomized Mixed-Methods Study

**DOI:** 10.64898/2026.08.24.26361252

**Authors:** Bryan J Bunning, Yingjie Weng, David JH Wu, Gavin Hui, Jessica E Hope, Ivan Lopez, Selin Everett, Vijay Pandurangan, Jonathan H Chen, Manisha Desai

## Abstract

Doctors increasingly rely on AI in the clinic, yet which report features make AI-generated responses useful and trustworthy remains unclear. In this randomized mixed-methods study, 34 oncology physicians provided 294 ratings of four blinded AI systems across five vignettes, alongside 20 semi-structured interviews analyzed with a prespecified LLM-assisted qualitative pipeline. Despite similar references, an evidence-graded report adapted from OpenEvidence was rated significantly lower in overall utility than standard OpenEvidence (mean difference, −0.96; 95% CI, −1.26 to −0.66; P<.001). Qualitative analysis identified six themes and seven design requirements. Oncologists valued rapid orientation, evidence retrieval, and verification, preferring concise, scannable reports with quantitative outcomes, recognizable bolded guidelines, explicit uncertainty, and verifiable citations. Trust deteriorated with citation mismatch, buried provenance, evidence misclassification, overconfident recommendations, and poor organization. Evidence presented differently can alter perceptions of clinical utility and trust; accuracy alone is insufficient, and report design must also be empirically evaluated.

## Introduction

Oncologists routinely make critical decisions amid rapidly changing guidelines, trials, and molecularly defined treatment options. Large language models (LLMs) have emerged as retrieval-and-summarization tools that can generate natural-language responses to open-ended clinical questions. Physicians are already using generative AI, with reports of more than 80% of physicians being regular users^1^. Physicians use both generalist and specialist models for clinical reasoning support and administrative work^2–6^. Oncology-specific surveys show similarly broad interest alongside concerns about misinformation, privacy, and overreliance^7^. In a small retrospective primary-care evaluation, OpenEvidence generated evidence-supported recommendations that aligned with physician plans and received high ratings for clarity, relevance, and evidence support; the investigators nevertheless emphasized the need for prospective evaluation in more complex and multidisciplinary settings^7,8^. These findings suggest that clinical use is advancing faster than our understanding of how clinicians evaluate, verify, and incorporate AI-generated reports, particularly in oncology, where the evidence base is rapidly evolving and decisions are high stakes^9^.

These systems support clinical questions by surfacing and organizing evidence; they should not be described as performing formal evidence synthesis. Nevertheless, evidence-synthesis frameworks provide useful principles for report design. Frameworks such as GRADE characterize certainty in a body of evidence according to domains including risk of bias, inconsistency, indirectness, imprecision, and publication bias^10,11^. LLMs are also being evaluated as tools for individual stages of formal evidence synthesis, including literature searching, screening, and data extraction^12–14^. Importantly, higher-order evidence appraisal remains challenging for LLMs: recent evaluations have identified limitations in risk-of-bias assessment and a tendency to insufficiently discount lower-quality evidence when synthesizing conclusions^15,16^. We hypothesize that making evidence characteristics and evidentiary strength more visible within AI-generated reports could improve their perceived utility. At the point of care, oncology teams are not conducting a systematic review, but they still need clear cues about study design, population relevance, clinical directness, and uncertainty. Presenting a phase III trial alongside an animal-model study without distinguishing their evidentiary roles can create a misleading impression that they provide equivalent support. Evidence strength and data provenance are complementary but distinct. Evidence hierarchy indicates how much confidence to place in a body of evidence, whereas provenance identifies the specific source material supporting a claim and enables direct verification by the physician user. HCI work on “traceable text”, in which generated claims link to supporting source passages, suggests that such interfaces can improve verification efficiency and accuracy, particularly when generated summaries contain errors^17,18^.

A key problem in clinical AI today is the recurring finding that physicians using AI perform worse than AI alone, suggesting that model capability is outpacing our ability to design effective physician–AI collaboration^19,20^. Model accuracy and medical knowledge alone also does not determine whether clinicians benefit^4,5^. In randomized diagnostic studies, access to a capable LLM did not necessarily improve physician performance, whereas a structured clinician–AI collaboration protocol improved performance and brought Physicians+AI to parity with AI alone^4,19^. These findings support evaluating the presentation and use of AI-generated evidence rather than treating model output as an isolated technical product to be benchmarked for accuracy. As we noted previously, “The [clinician-AI collaboration] study provides evidence of unrealized opportunities at the intersection of design, engineering, and medicine for enhancing clinician-AI collaboration. Additional cross-disciplinary research and methods development are needed to realize… the possibilities^19^.” Accordingly, clinical AI evaluations should consider report and physician-AI interaction design, not only the accuracy of the underlying model^21^. There exist design flaws which inflict distrust between the Physician user and the AI response. What are those gaps? What features may improve trust and collaboration?

The primary objective of this study was to elicit drivers of oncologist trust and key features of AI-generated literature review reports via blinded ratings and semi-structured interviews to improve upon physician-AI collaboration. We hypothesized that a modified report may improve collaboration by focusing on the strength of evidence reported. Through a detailed prompt we reorganized the response and citations returned by a widely used clinical AI platform to make study design and evidentiary strength more visible, and rank order the references by its strength of evidence, emphasizing large scale clinical trials. We then compared it with reports from clinical and general-purpose AI systems in a randomized mixed-methods study. Quantitative ratings assessed perceived utility and report attributes, while semistructured interviews and free-text responses identified the presentation features, verification practices, workflow needs, and trust concerns that clinicians considered most important. A secondary objective was to demonstrate a prespecified LLM-assisted qualitative-analysis workflow designed to support consistent, auditable extraction of opinions with investigator oversight. The study therefore used report comparisons to generate design evidence, rather than as a product-ranking exercise or a test of whether one model was clinically correct.

## Results

### Participant characteristics

The study design and flow are shown in Figure 1. 34 clinicians participated across 20 interview sessions in March-April 2026 assessing 5 oncology vignettes and associated model responses from 4 AI systems. Every participant completed at least one quantitative rating and contributed qualitative data through a recorded interview and/or free-text responses; therefore, as prespecified, both the quantitative and qualitative analysis sets included the total 34 participants. Participants were recruited across multiple institutions, with the largest groups from Stanford (16/34, 47%) and UCLA (8/34, 24%) (Table 1). Most participants were oncology fellows (18/34, 53%); residents and attending/faculty physicians each comprised 8 participants (24%). The median years in practice, including fellowship years, was 2.5 (IQR, 3.1). Oncology focus areas included radiation oncology (13/34, 38%), pediatric hematology-oncology (10/34, 29%), medical oncology (6/34, 18%), and hematology-oncology (5/34, 15%).

**Figure 1:**
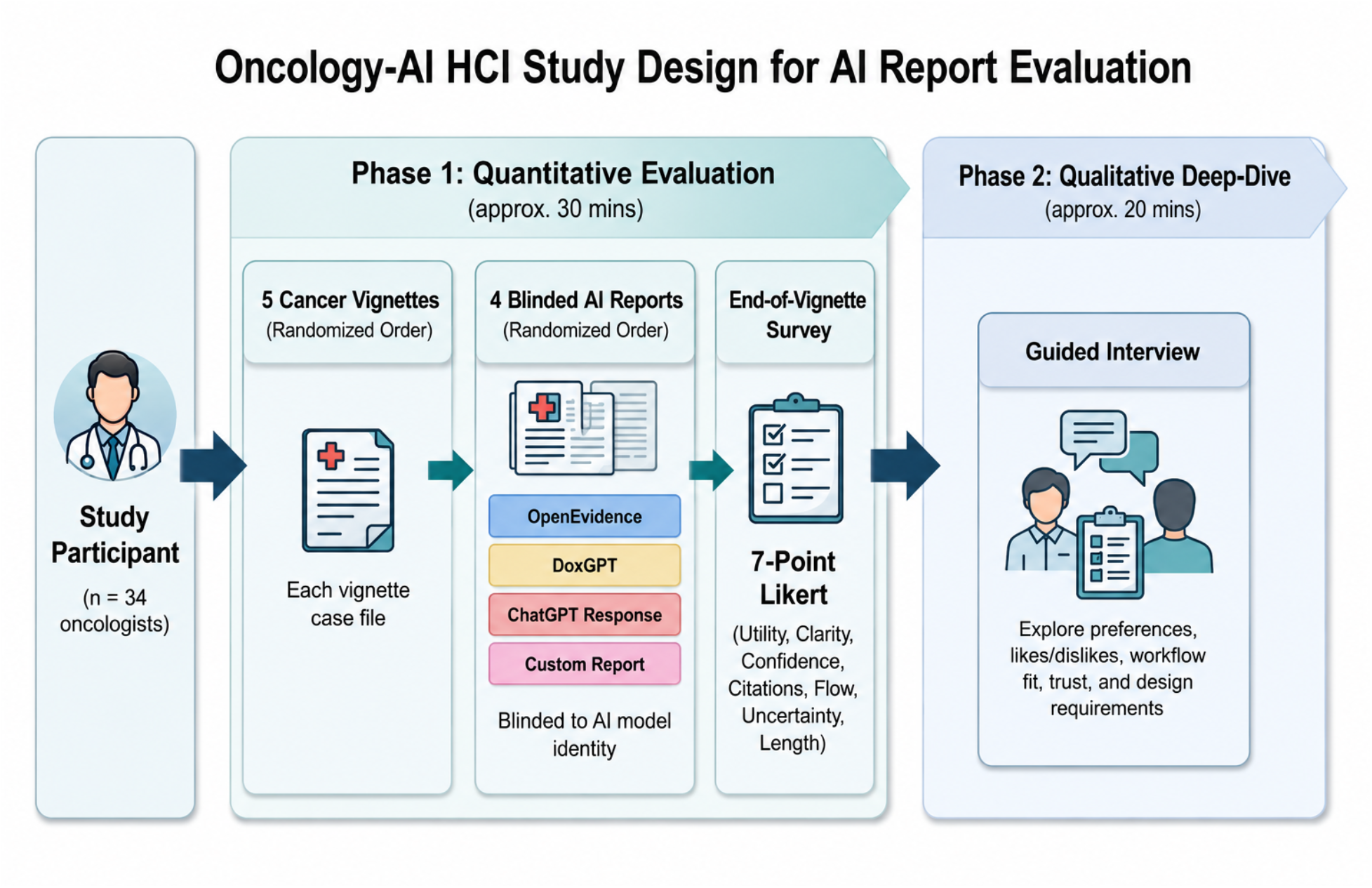
Study design and flow. After brief instructions, participants completed as many randomized vignette reports as they were able in 60 minutes. Vignette order was randomized, and then within each randomized vignette, the ordering of AI reports (all generated March 3 2026) were further randomized. A participant was provided all 4 AI reports for each vignette before being assigned a new vignette. The qualitative deep dive was a semi-structured interview, starting with a few uniform questions (see supplement).

**Table 1:** Participant demographics. Self-reported demographics and AI use of the participant population. Due to unanswered questions in the survey, the denominators for AI use frequency (n=33) and use of different AI toolings (n=32) differ from the total population (n=34).

| Characteristic | N = 34 |
| --- | --- |
| **Institution** |  |
| Stanford | 16 (47%) |
| UCLA | 8 (24%) |
| Medical College of Wisconsin | 2 (5.9%) |
| Brooke Army Medical Center | 1 (2.9%) |
| Cone Health | 1 (2.9%) |
| Mayo Clinic | 1 (2.9%) |
| MD Anderson Cancer Center | 1 (2.9%) |
| Memorial Sloan Kettering cancer center | 1 (2.9%) |
| NYU Langone Health | 1 (2.9%) |
| Roswell Park Comprehensive Cancer Institute | 1 (2.9%) |
| University of Louisville | 1 (2.9%) |
| **Training level** |  |
| Resident | 8 (24%) |
| Fellow | 18 (53%) |
| Attending/Faculty | 8 (24%) |
| Years in practice, median (IQR) | 2.5 (3.1) |
| **Oncology focus** |  |
| Hematology Oncology | 5 (15%) |
| Medical Oncology | 6 (17.9%) |
| Pediatric Hematology Oncology | 10 (29%) |
| Radiation Oncology | 13 (38%) |
| **AI use frequency** |  |
| Never | 1 (3.0%) |
| Barely | 5 (15%) |
| Sometimes | 11 (33%) |
| About half the time | 2 (6.1%) |
| Consistently as a case warrants it | 6 (18%) |
| Most of the time | 5 (15%) |
| Always | 3 (9.1%) |
| Uses ChatGPT | 22 (69%) |
| Uses OpenEvidence | 27 (84%) |
| Uses DoximityGPT | 4 (13%) |
| Uses UpToDate AI | 7 (22%) |
| Uses Claude | 7 (22%) |
| Uses Perplexity | 4 (13%) |
| Uses NotebookLM | 3 (9.4%) |

Participants reported substantial prior exposure to AI tools. OpenEvidence was the most commonly used tool (27 participants, 84%), followed by ChatGPT (22 participants, 69%). Smaller proportions reported using UpToDate AI or Claude (7 participants each, 22%), DoximityGPT or Perplexity (4 participants each, 13%), and NotebookLM (3 participants, 9.4%). Most participants described some degree of AI use in clinical or clinically adjacent contexts, although reported use frequency and risk tolerance varied (Table S1).

### Qualitative findings

A figure showcasing the prespecified qualitative analysis pipeline is shown (Fig 2). As part of our prompt analysis, the AI generated synthesis of interviews included multiple pre-defined components (supplement, fig S2). Qualitative findings were synthesized across 20 interviews, all of which contained usable data across 5 hours 59 minutes and 46 seconds of total interview time. All participants verbally consented to recording of the qualitative interviews. Due to scheduling, some interview sessions contained more than one participant. The LLM based pipeline structured data into a qualitative knowledge base, which identified six cross-interview themes, five synthesized likes, five synthesized dislikes, and seven design requirements, alongside 19 workflow-related findings, 16 trust and safety findings, four key tensions or tradeoffs, and three minority or divergent perspectives (Fig S2, S4). The six overarching themes were supported by 13 to 18 interviews each (65%–90%), with the greatest convergence around the importance of visible, clinically interpretable evidence (18/20, 90%) and brief, scannable reports suited to time-constrained clinical workflows (17/20, 85%). The final synthesis included 32 representative or example quotations. Overall, the findings reflected strong agreement about the criteria clinicians used to evaluate AI-generated literature reports, while revealing heterogeneity in preferred report format, level of detail, and degree of recommendation-forward language (Fig 3, Supplement). The complete structured knowledge base file is publicly available.

**Figure 2.**
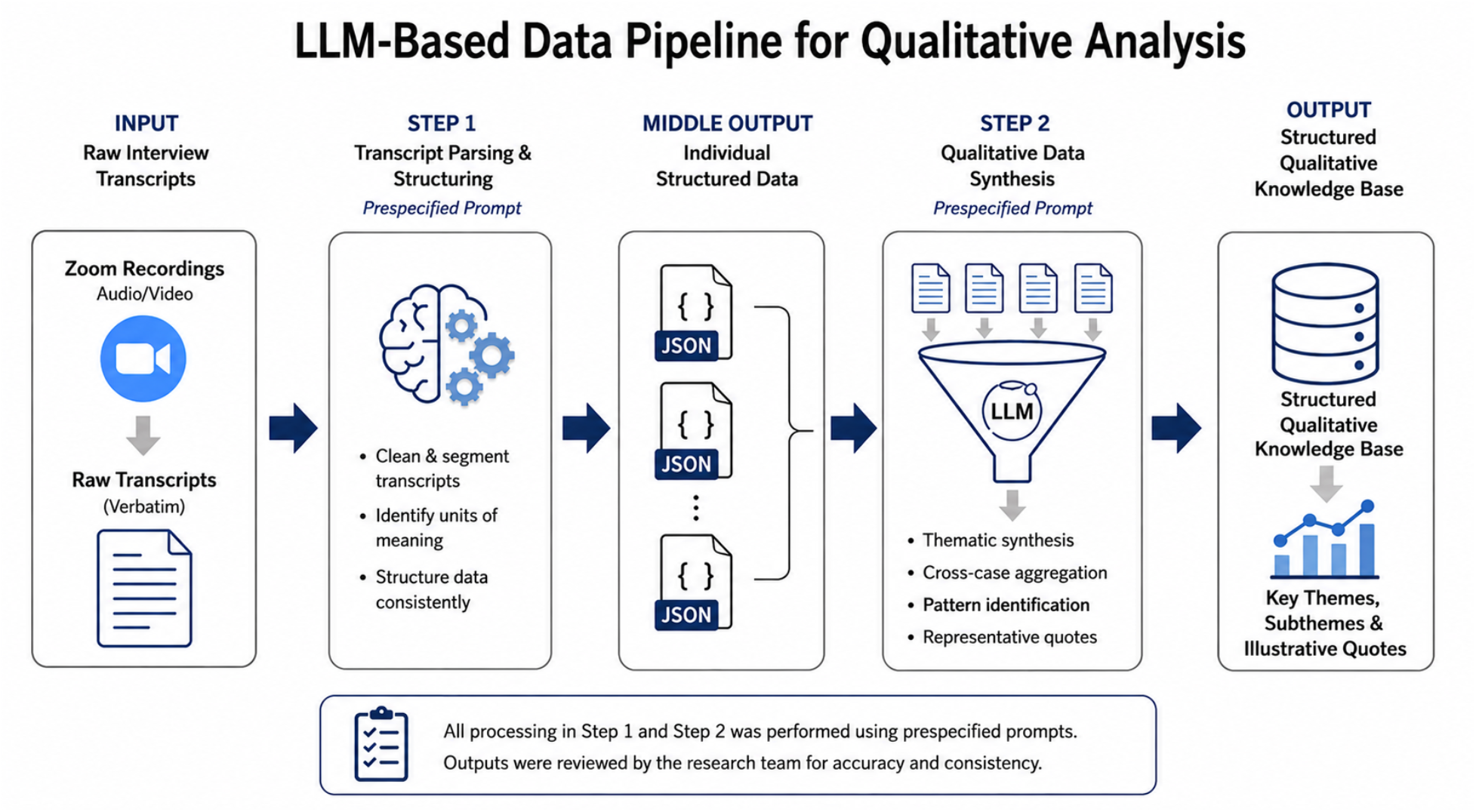
Methodology for qualitative assessment and analysis. Methodology for qualitative assessment and analysis of 20 oncologist semi-structured interviews. Each deidentified transcript was analyzed using a prespecified prompt and converted into a structured JavaScript Object Notation (JSON) file, a machine-readable format that organized participant-derived themes, preferences, workflow observations, trust and safety findings, and representative quotations into consistent fields. The 20 interview-level JSON files were then synthesized into a single study-level structured knowledge base that summarized cross-interview themes and insights while preserving supporting quotations and their source interviews.

**Figure 3.**
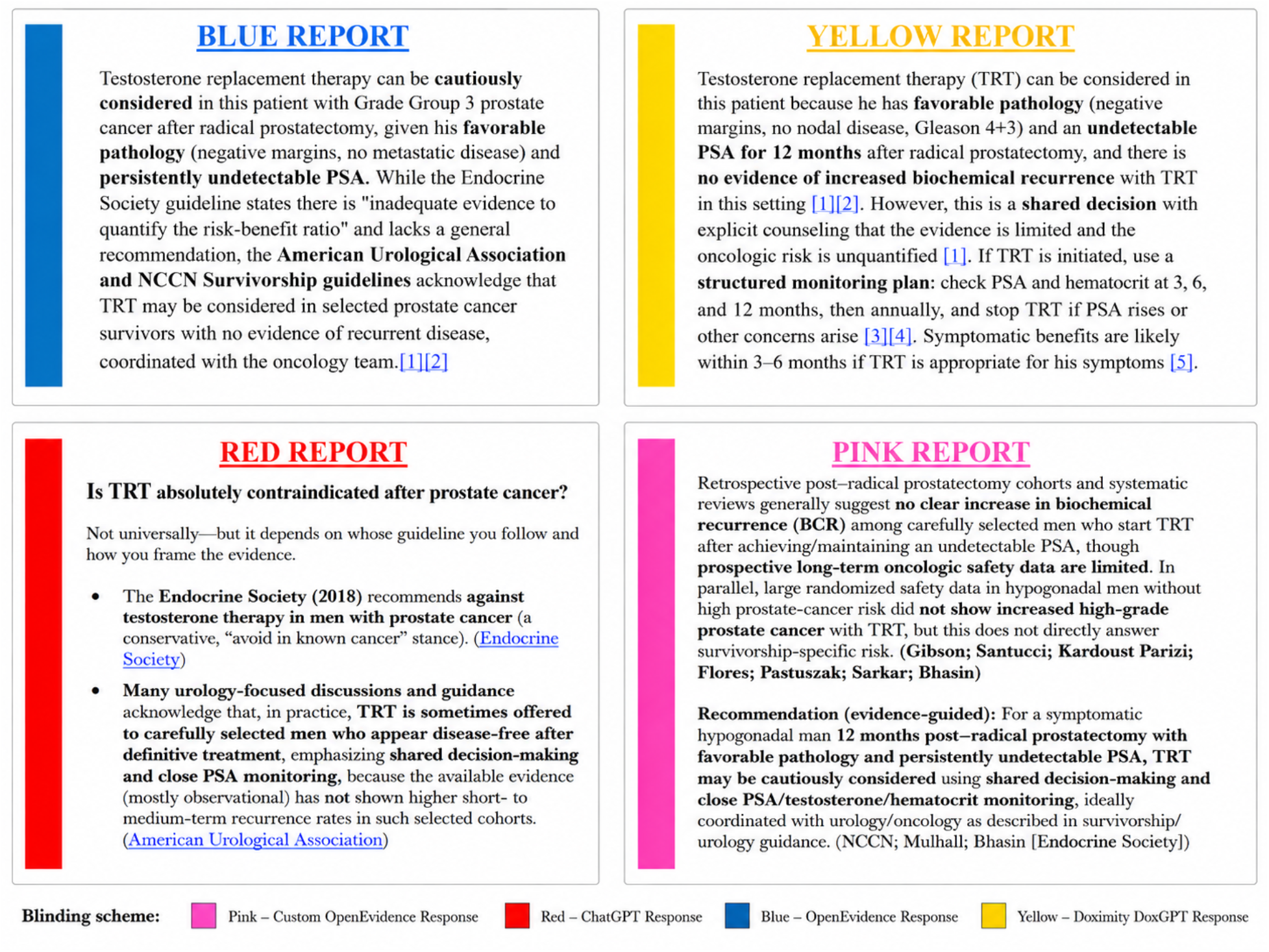
Example text of AI reports shows heterogeneity in design. A selected snippet of AI responses to an identical vignette, concerning the risk-benefit of hormone replacement therapy after radical prostatectomy in prostate cancer, is shown. There exists heterogeneity in the structure, ordering, and bolding decisions between models. In this report, all four AI models state a similar sentiment, and use similar underlying references (NCCN, Endocrine Society, AUA guidelines), yet differ in the decisions to describe or visualize it. Of note, OpenEvidence (Blue Report), has the least bolding in this snippet, and elects to bold the guideline source or societies it is referring to. The OpenEvidence model chose to bold the AUA and NCCN, but not the Endocrine Society. All model responses occurred on March 3, 2026.

Across 20 interview sessions inclusive of all 34 participants, clinicians consistently emphasized design requirements of AI output. AI-generated oncology literature reports should be concise, scannable, and easy to verify (Table 2). The most frequently supported design requirements were a brief, top-loaded clinical summary (7 interviews), direct presentation of clinically relevant study-level evidence such as key endpoints and safety data (6 interviews), and accurate source traceability with clearly labeled (National Comprehensive Cancer Network, American Society of Clinical Oncology, etc) and clickable citations (5 interviews). Participants also favored explicit communication of uncertainty, structured formatting, and the ability to adjust report depth to the clinical task. Interestingly, physicians also asked for explicit separation between literature review and AI reasoning/recommendation.

**Table 2.** Design requirements and mapped preferences for AI-generated oncology literature-review reports. Counts reflect the number of interviews containing supporting evidence rather than the number of individual mentions; percentages use all 20 interviews as the denominator. Because interviews were semistructured and participant-led, the requirements and preference signals emerged spontaneously rather than in response to a standardized design question. Explicitly stated synthesized likes and dislikes were mapped post hoc to an aligned relevant design requirement. As the design requirements and stated likes and dislikes were separate questions in the interview, their counts may differ. Quotes are representative verbatim excerpts selected from the synthesis.

| Summary | Design requirement | n (%) | Explicitly stated Likes/Dislikes | Why it matters | Risk if ignored | Representative quote |
| --- | --- | --- | --- | --- | --- | --- |
| Brief, top-loaded summary | Lead with a brief, top-loaded clinical summary or bottom line that can be read in seconds, with deeper evidence available underneath. | 7 (35%) | liked concise bottom lines (9/20); disliked long or repetitive reports (n=9). | Participants repeatedly described between-patient use, clinic time pressure, and low tolerance for long reading. A front-loaded summary was one of the clearest adoption conditions across interviews. | The tool is likely to be skipped in live workflow because the reading burden will exceed perceived benefit. | "one page max... bite-sized, easy to digest." |
| Include study level endpoints | Show clinically relevant study-level evidence directly in the report, including trial phase/type, patient population, key outcomes, and toxicity or adverse-event data where relevant. | 6 (30%) | liked visible quantitative trial outcomes (n=9). | Participants did not want conclusions alone; they wanted enough quantitative detail to judge whether the evidence actually supported the answer. | Clinicians may view the output as superficial or untrustworthy and revert to manual literature search. | "I want to know the outcomes... the percentages, like, PFS benefit." |
| Source Traceability | Provide direct, accurate, and well-labeled source traceability, ideally with clickable links to the exact guideline, paper, paragraph, table, or figure supporting each claim. | 5 (25%) | liked recognizable guideline anchoring (n=8/20); disliked citation mismatch or weak traceability (n=4). | Traceability was a core trust driver and a common verification behavior. Participants wanted to inspect source support quickly without repeating the entire search process. | Trust will erode quickly, especially after even a small number of citation mismatches or unlabeled links. | "I, like, can't find what it said in the citation." |
| Separate evidence from AI recommendation | Separate evidence presentation from recommendation language and communicate uncertainty explicitly when the evidence is limited, mixed, or context-dependent. | 5 (25%) | disliked overconfident or directive recommendations (n=4). | Participants were often comfortable with summarized evidence but much less comfortable with unsupported decisiveness. Calibrated uncertainty was treated as a safety feature. | Overconfident summaries may mislead clinicians, especially in nuanced cases, and can poison trust in the entire report. | "I really appreciate the use of them for summarizing information, but I do not really look to these tools for recommendations about what to do." |
| Easy to read formatting | Use sectioned, scannable formatting with bullets and selective tables, but avoid dense paragraph blocks, crowded reference dumps, and tables that consume space without adding signal. | 5 (25%) | liked predictable clinical sections (n=4); disliked crowded or visually noisy layouts (n=5). | Participants agreed that structure matters, but table preferences varied. The cross-case pattern favored readability over any one fixed layout. | Even accurate content may be ignored if clinicians must work too hard to parse it. | "definitely putting things into different sections is helpful." |
| Adaptable context depth | Allow output depth or format to adapt to task and user, such as quick clinic view versus deeper pre-visit or academic view. | 5 (25%) | liked patient- and task-specific framing (n=4). | Participants described different needs for clinic, studying, note support, literature scouting, and unfamiliar topics. Several explicitly said that one report was not enough or that the ideal output would combine multiple styles. | A fixed format may fit one workflow moment while failing another, limiting adoption breadth. | "the optimal one for me would be giving me a direct answer... then a small summary table." |
| No AI tone | Avoid chatty AI tone, decorative parentheticals, and interpretive prose that sounds more like generated commentary than clinical evidence support. | 3 (15%) | disliked AI-sounding or editorialized tone (n=4). | Tone itself affected acceptance and credibility for several participants, independent of the underlying content. | The report may be dismissed as unserious or less trustworthy even when it contains relevant information. | "the writing style was very AI-focused, which is... a big turnoff." |

Overall preferences similarly favored concise bottom-line summaries and quantitative trial outcomes, each supported by 9 interviews, followed by guideline anchoring and visible evidence hierarchy, supported by 8. Common dislikes included reports that were overly long or repetitive (9 interviews), visually crowded or difficult to parse (5), and overly confident, editorialized, or poorly traceable (4 each). Trust was strengthened by recognizable guideline sources, numerical outcomes, and accurate citations, whereas citation mismatch, evidence misclassification, and overconfident recommendations were recurrent trust and safety concerns (Table S2). Verbatim topline results pulled directly from the structured qualitative knowledge base without edit are included in the supplement.

### Quantitative Outcomes

Across the 34 clinical experts, ratings were balanced across patient cases (n=54, 55, 75, 52, and 58) and report types (OpenEvidence, n=75; ChatGPT, n=73; DoximityGPT, n=70; modified OpenEvidence, n=76). Mean perceived overall utility across report types was 4.5 on the 7-point likert scale. In the prespecified analysis, using a mixed-effects model, the modified OpenEvidence report had lower perceived utility than the original OpenEvidence report (mean difference, -0.96; 95% CI, -1.26 to -0.66; P<.001), after accounting for clustering by participant and vignette. ChatGPT did not differ from OpenEvidence (mean difference, -0.02; 95% CI, -0.32 to 0.29); nor did DoximityGPT (mean difference, 0.11; 95% CI, -0.20 to 0.42) (Figure 4).

**Figure 4:**
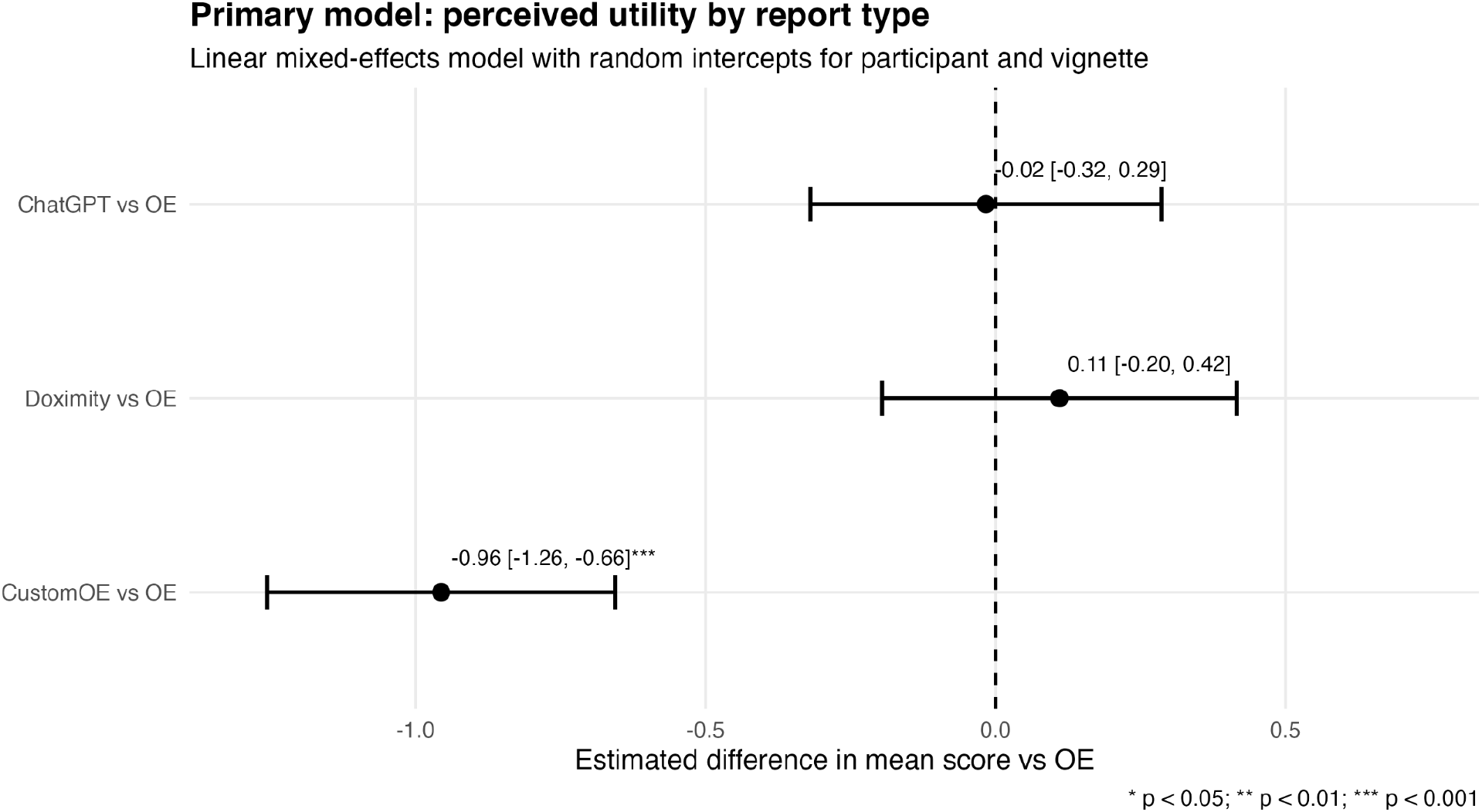
Oncologist perceived utility by report type on a 7-point likert scale. The prespecified endpoint comparing the OpenEvidence to the custom prompt derived from the OpenEvidence response failed, as we hypothesized the custom report would be better perceived. The OE and CustomOE models were derived from the same set of references/citations, suggesting design can differentially affect perceptions of quality. A linear mixed-effects model with random intercepts for participant and vignette was employed to adjust for clustering. This model is derived from 294 participant-vignette-report scores from 34 participants.

Differences by report type varied across report attributes (Fig S1). Compared with standard OpenEvidence, ChatGPT had significantly lower citation quality ratings (mean difference, -0.83; 95% CI, -1.18 to -0.48), but did not differ meaningfully on clarity, confidence in action, communication of uncertainty, order and flow, or length. DoximityGPT also had lower citation quality ratings than standard OpenEvidence (mean difference, -0.63; 95% CI, -0.98 to -0.28), while receiving improved ratings for length (mean difference, 0.51; 95% CI, 0.10 to 0.92). Other DoximityGPT subcategory ratings were not significantly different from standard OpenEvidence.

The customized OpenEvidence-style report was rated lower than standard OpenEvidence across all six subcategories. The largest deficits were observed for communication of uncertainty (mean difference, -1.10; 95% CI, -1.43 to -0.78), order and flow (mean difference, - 1.04; 95% CI, -1.39 to -0.68), clarity (mean difference, -1.03; 95% CI, -1.33 to -0.74), and confidence in action (mean difference, -0.99; 95% CI, -1.30 to -0.68). The customized report also had lower citation quality ratings (mean difference, -0.78; 95% CI, -1.12 to -0.43) and lower length ratings (mean difference, -0.43; 95% CI, -0.84 to -0.03).

In the survey’s overall ’best report’ question, the distribution of selections did not differ significantly from a uniform 25% distribution (P=.14). In the overall ’worst report’ question, the distribution did differ from uniform, with the modified report selected most frequently (p=.01, n=4 unanswered). Most participants preferred reports 1-2 pages long (53.3%), followed by 1 page (26.7%), 2-3 pages (10.0%), less than half a page (6.7%), and more than 3 pages (3.3%) (Fig S3).

## Discussion

In this mixed-methods evaluation of AI-generated oncology literature-review reports, perceived clinical utility was influenced by tone, structure, and design features of the AI responses, which had heterogeneity across the models in the study (Fig 4). Workflow fit, response format, data provenance, named medical society guideline positions, and trust-enabling design decisions appear central to whether clinicians perceive an AI-generated report as useful. Qualitative findings clarified a pattern: oncologists valued AI outputs primarily as tools for rapid orientation, evidence retrieval, and verification rather than as autonomous decision-makers. These results suggest that for clinical AI to provide value to physicians, content accuracy is necessary but not sufficient. This interpretation is consistent with a recent systematic review and meta-analysis showing that human–AI combinations often fail to outperform the better-performing member alone, with the largest deficits observed in decision-making tasks, underscoring that effective collaboration depends on how AI is integrated into human workflows rather than on model capability alone^22^.

We hypothesize clinician trust of AI is associated with how evidence is displayed and communicated, not just in an AI response’s objective accuracy. Theoretically, on the assumption the medical content from the AI is correct, what is the optimal way to convey the content to an Oncologist? Participating physicians repeatedly emphasized that recommendations were more credible when reports made the source of evidence visible, recognizable, and easy to verify. This is directly supported by the significant difference between OpenEvidence and our customized OpenEvidence reports, with the custom report performing worse, which included similar source data and citations. The customized report was derived entirely from the OpenEvidence output with a prompt asked to change the ordering of information by ranking the references and the strength of study tied to each reference. We believe the aesthetics and structure of the custom report decreased its perceived utility. The lower rating for the modified report was treated as design evidence to refine future reports, rather than as failure of a model-performance. In our data, design features such as naming guidelines or major trials, bolding the source or society of a recommendation in the text, providing direct hyperlinks with highlighted sections within the reference, and surfacing study-level details appeared to distinguish more trusted reports from less trusted ones. This finding is consistent with data provenance and traceability literature as well as emerging evaluations specifically of clinical AI tools showing that physician judgments depend not only on answer quality, but also on source quality, verifiability, and clinical utility^2^.

More broadly, these results highlight that form implies function: the layout, hierarchy, and reference structure of an AI output may shape whether clinicians treat it as a useful evidence-review assistant or as an unsupported recommendation engine. Physicians may remain appropriately skeptical of “AI answers,” while still welcoming AI systems that retrieve, organize, and contextualize guidelines, trials, and citations. Given this finding, it may be prudent to clearly separate the literature review section of AI responses from the AI’s interpretation or recommendation. Our data is supportive of the theory that aspects of trust can be improved with UI and design decisions which, counterintuitively, may be independent of the clinical answer’s accuracy itself. This resembles broader evidence that presentation cues can influence perceived expertise; for example, physician attire affects patient judgements of professionalism and experience, the so-called “white coat effect”^23^. Analogous interface cues may shape perceptions of clinical AI even when underlying content is unchanged, underscoring the need to distinguish warranted trust from presentation-driven trust. An AI response must likewise be “dressed up” for physician use. We aggregate our findings of what a good and bad report may look like, according to our data, in Figure 5.

**Figure 5.**
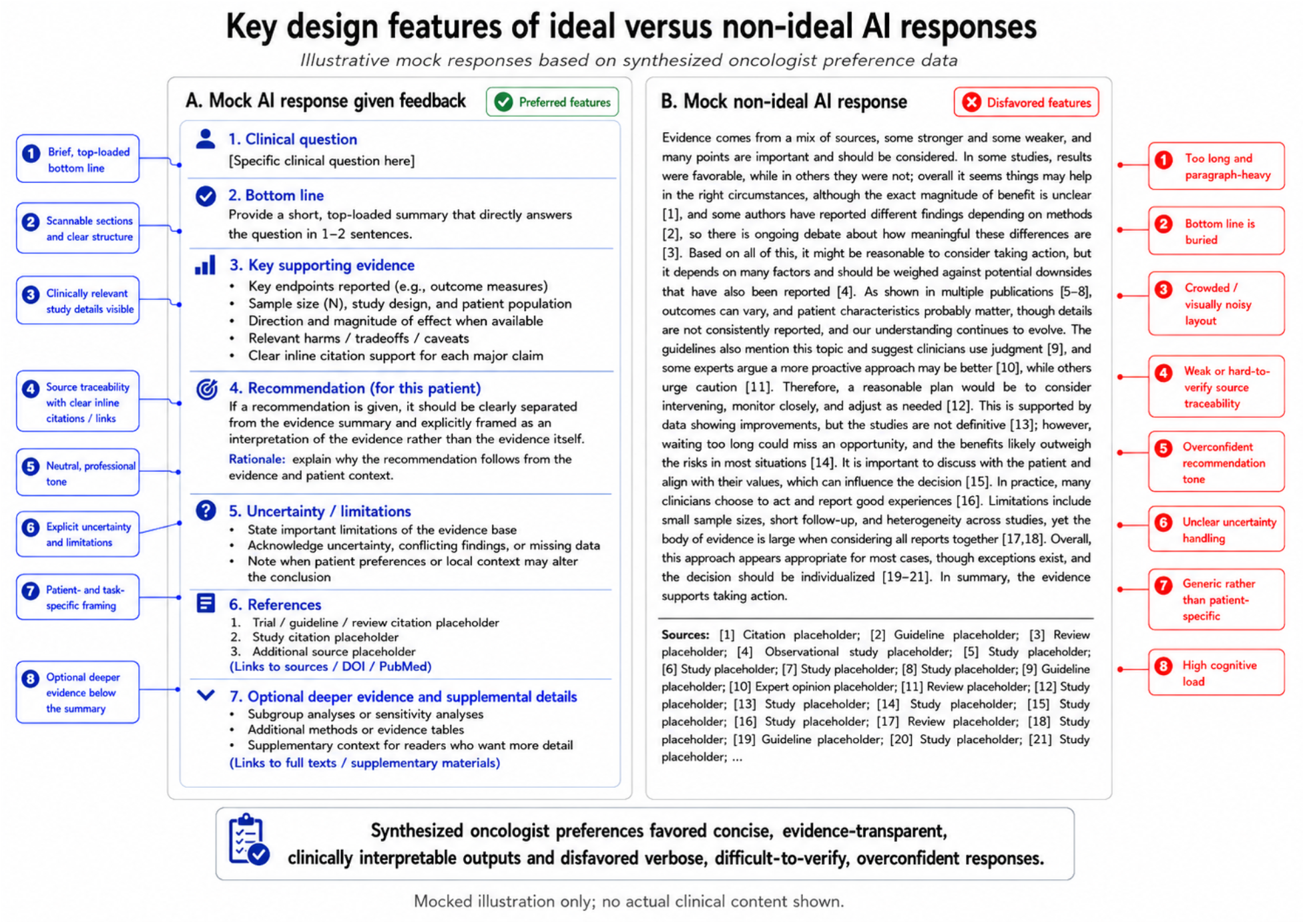
Integration of synthesized oncologist preferences into an illustrative AI response design. Key findings from the cross-interview qualitative synthesis were integrated into contrasting mock examples of an AI response incorporating participant feedback and a non-ideal AI response. Main findings include the intentional separation of key evidence from the literature from an “AI recommendation”, improvements in source traceability, and the inclusion of additional clinically relevant study details for clinical trials referenced in a response.

These findings also highlight an underdiscussed design problem in clinical AI evaluation. Model benchmarking often emphasizes correctness, but real clinical adoption depends on whether the system is trusted by physicians and supports (not automates!) accountable, verifiable clinical reasoning^5,24^. Recent independent benchmarking found that frontier general-purpose LLMs outperformed specialized clinical AI tools, including OpenEvidence, on medical knowledge benchmarks and blinded clinician ratings of real world queries; notably, that evaluation did not assess citation quality. Our findings therefore highlight a complementary dimension of clinical AI evaluation: answer performance alone does not establish whether evidence is presented in a form clinicians can efficiently interpret, verify, and appropriately trust. Being right is not enough. The models assessed in this study showed great heterogeneity in their design decisions in responses. Further, in oncology, where recommendations often depend on disease subtype, treatment line, molecular features, trial eligibility, evolving guidelines, and patient preference, citation quality is not solely a cosmetic feature. It is a core component of clinical usefulness. Reports that rely on recognizable guidelines, pivotal trials, and clinically relevant outcomes such as progression-free survival, overall survival, toxicity, and subgroup effects may generate more appropriate trust than reports that cite weaker evidence, bury provenance, or present overconfident interpretations which may in turn degrade trust^25^.

Standard OpenEvidence performed similarly to ChatGPT and DoximityGPT on overall utility and outperformed both on citation quality, while the customized OpenEvidence-style report was rated significantly lower than standard OpenEvidence across overall utility and all measured subdomains. As our custom report was significantly worse, not better, than OpenEvidence, our primary endpoint failed. We had hypothesized that by reranking responses by the strength of evidence, physicians would find the reports more useful and concise. The subdomain pattern suggests that foregrounding evidence hierarchy in the design we chose came at a usability cost. The customized report was perceived as less clear, less well organized, worse at communicating uncertainty, and less concise. Because the intervention altered multiple presentation features simultaneously, the study cannot attribute the lower utility rating to any single design choice. While our attempt at a custom report to improve these perceived qualities failed, the thesis that physicians wanted the strength of evidence incorporated into AI responses is supported by qualitative interview data, as well as externally to this study in OpenEvidence’s recent release of EvidenceGrade™; evidence-strength display is an active product design priority^26^.

A secondary contribution of this study is methodological. We used prespecified language-model prompts and structured JSON outputs to efficiently aggregate qualitative physician preferences across interviews and free-text responses. Emerging work on LLM-assisted qualitative analysis suggests that analytic performance depends not only on the underlying model, but also on the structure of the workflow, including decomposition of analysis into discrete stages and preservation of intermediate outputs that can be inspected and systematically aggregated^27,28^. Our approach similarly used a schema-constrained JSON representation as an intermediate analytic layer between source text and study-level synthesis. Rather than moving directly from individual responses to narrative conclusions, this intermediate structure consistently captured themes, likes, dislikes, workflow constraints, trust and safety considerations, design requirements, and supporting evidence before cross-participant aggregation. This approach provides a machine-readable, auditable, and sharable analytic record, facilitates consistent synthesis across heterogeneous qualitative data, and reduces reliance on unstructured post hoc interpretation. The method is model agnostic and may improve as future models are deployed, while the structured intermediate outputs remain available for review, reanalysis, or application of alternative models. Importantly, however, the flexibility of language-model pipelines also introduces substantial analytic degrees of freedom. Prespecification of prompts, output schemas, and analytic procedures is therefore essential to reduce the risk that investigators iteratively adapt an AI pipeline to favor a particular finding or interpretation. Prespecification should consequently be viewed as a central safeguard against post hoc selective analysis and cherry-picking in LLM-assisted qualitative research.

This study has several limitations. First, product outputs are time-sensitive: AI tools, retrieval systems, model versions, and interface designs change rapidly, meaning that these findings should be interpreted as evidence about design principles and clinician preferences rather than rankings of specific products. Second, it evaluated perceived utility, workflow fit, and trust-related design features, not medical correctness or patient outcomes. The reports studied here should therefore be interpreted as evidence-review aids rather than autonomous clinical decision systems. Third, the sample was drawn largely from academic oncology settings, which may limit generalizability to community practice, non-oncology specialties, or clinicians with different levels of AI familiarity. Fourth, oncologist subspecialties were not always matched to vignette content, which may have influenced ratings of utility and confidence. Fifth, because sessions were capped at approximately one hour, not all participants completed all vignettes. Sixth, the tone and response of certain AI models are quite distinct, which could have contributed to accidental unblinding to power users of the models.

Future work should evaluate whether trust-enabling design features causally improve clinical use, verification behavior, and decision quality. Randomized studies could isolate specific interface elements, such as visible guideline anchoring, bolded evidence sources, direct citation links, evidence-level labels, uncertainty statements, and expandable study details. Additional studies should assess whether these features reduce inappropriate reliance while preserving efficiency gains, particularly among trainees or clinicians working outside their subspecialty. Methodologically, the use of multiple model providers may provide additional robustness towards an LLM based summarization of interviews. Additionally, a blinded comparison between an LLM-derived qualitative analysis and a human qualitative analysis is warranted. Ultimately, clinical AI tools will be most valuable when they combine strong medical intelligence with transparent evidence display, workflow-aligned formatting, and design choices that preserve clinician control over interpretation and action.

Though models, companies, and medical AI products will inevitably change, we believe that oncologists’ preferences in design are more stable and immutable given the inherent nature of human taste. “Beauty is in the eye of the beholder”, but through these multiple semi-structured interviews with oncologists we have surfaced that there are shared and aligned design principles that nurture trust-building by their physician-users and likewise, absence of these principles can damage user-trust especially in a high-stakes setting such as oncologic consultation. The optimal clinician-using tool will not look like the optimal completely autonomous clinical AI system. Development and advances in human-computer interaction with LLMs is needed to realize the potential of AI in clinical decision support.

## Methods

### Study design and oversight

We conducted a remote, within-participant, randomized vignette study evaluating clinician perceptions of AI-generated oncology literature-review reports. The study was designed as a mixed-methods evaluation of perceived report utility, preference, workflow fit, and opportunities for improving AI-assisted literature synthesis in oncology. Each participant reviewed clinical vignettes describing oncology patient cases paired with multiple blinded AI-generated literature-review reports and completed structured ratings, comparative preference questions, free-text responses, and a post-task interview when consent for recording was provided.

The LLM prompts and initial statistical analysis plan were preregistered on the Open Science Framework prior to data collection. The finalized statistical analysis plan was preregistered prior to unblinding and outcome analysis. The study was reviewed by the Stanford Institutional Review Board and determined to be exempt status human subjects research under protocol 85187. All participants provided informed consent before study participation. The study was conducted remotely using Qualtrics for survey administration and Zoom for optional recorded exit interviews.

### Participants

Eligible participants were physicians with oncology expertise who had completed at least postgraduate year 2 training, were fluent in English, were able to provide informed consent, had access to the internet and a computer for remote participation, and were available for an approximately 1-hour study session. The study preferentially recruited physicians with specialty oncology training or current oncology practice experience, including hematology-oncology, medical oncology, radiation oncology, and related oncology-focused clinical roles. Participants who were not fluent in English were excluded.

Participants were recruited from a combination of professional networks, email lists, and cold outreach throughout February-April 2026. Recruitment included outreach from investigators at their local institutions as well as cold outreach to an informal whatsapp group of residents and attendings created to facilitate collaboration from the American Society for Radiation Oncology (ASTRO) conference. Interviews occurred from March-April 2026. We targeted enrollment of 30 clinicians to allow for attrition and incomplete vignette completion while preserving precision for the primary within-participant comparison. For their time, physicians were compensated based on their level of expertise: $100 for residents, $150 for fellows, and $199 for attending physicians. Per IRB approval, participants were explicitly asked if they were interested in having their qualitative interview recorded for research purposes.

### Study materials

The study materials consisted of five oncology literature-review vignettes, each posing a clinical question that required evidence synthesis rather than autonomous medical decision-making. Each vignette mimics a plausible scenario for an oncologist, but includes no real patient data. For each vignette, we generated four reports: (1) OpenEvidence, (2) a customized report produced with GPT-5.2 Thinking using the OpenEvidence report as the sole source, (3) ChatGPT using GPT-5.2 Thinking, and (4) DoxGPT. OpenEvidence and DoxGPT were included as clinician-facing medical AI tools, whereas ChatGPT represented a widely used general-purpose LLM. The same verbatim vignette was submitted to each system on the same date: March 3 2026. Reports were presented under blinded labels. The study evaluated perceived utility and design; it did not assess or attempt to equalize clinical accuracy across reports.

The customized report was designed to foreground the hierarchy and strength of evidence by prioritizing guidelines and large clinical trials and deemphasizing preclinical studies and small observational cohorts. The model was instructed to reorganize, not supplement, the OpenEvidence output; it could use only the source report and was prompted not to introduce outside information. The prompt and comparison of references between the Open Evidence report and the custom report is provided in the Supplement.

All AI reports were generated before participant review on March 3rd 2026 using the verbatim vignettes as input. Prompts, source outputs, and representative vignettes are provided in the supplement and public repository.

### Randomization and masking

Randomization occurred separately for each participant. Qualtrics first randomized the order of the five vignettes and then randomized the order of the four blinded reports within each vignette. Each report source was assigned a color label that remained consistent across vignettes for that participant. The study team member (B.B.) administering the session remained blinded to report identity and order unless a participant explicitly identified a source. Prespecified report mappings were applied after data export.

Because each participant could rate multiple reports across multiple vignettes, the study design generated repeated observations clustered within participants and vignettes. The survey was capped at approximately 1 hour, so not all participants were expected to complete all vignettes.

### Study procedure

After consent, participants completed baseline questions including training level, institution or practice setting, and additional demographic or professional characteristics when collected. Participants then reviewed randomized oncology vignettes and the blinded AI-generated reports associated with each vignette. Immediately after reviewing each report, participants rated its overall utility and several prespecified subdomains on 7-point Likert-type scales. Participants also provided free-text comments about report strengths, weaknesses, and improvement opportunities.

At the end of the survey, participants selected the best and worst report type overall and indicated their preferred or optimal report length. Participants then completed an optional semistructured exit interview focused on current AI-tool use, perceived utility of the reports, trust and verification behaviors, workflow fit, friction points, and recommendations for improving AI-generated oncology literature reviews (see supplement). Interviews were audio-recorded only with participant permission and transcribed using Zoom’s embedded tooling for qualitative analysis.

### Outcomes

The primary outcome was participant-rated overall utility of each vignette-specific AI-generated report, measured on a 7-point Likert-type scale and coded numerically from 1 to 7. The primary prespecified comparison was the difference in mean overall utility between the customized OpenEvidence-style report and standard OpenEvidence.

Secondary quantitative outcomes included utility contrasts between each additional report type and standard OpenEvidence; six 7-point Likert-type subcategory ratings evaluating report features including: clarity, confidence in action, citation quality, order and flow, communication of uncertainty, and length. Additional questions included participant selection of the best report type; participant selection of the worst report type; and participant-reported preferred report length. Qualitative outcomes included inductively derived themes, likes, dislikes, workflow constraints, trust and safety considerations, and actionable design recommendations from free-text comments and interview transcripts described below.

### Quantitative statistical analysis

The full analysis set included all consented participants who began the Qualtrics instrument and provided at least one overall utility rating. The qualitative analysis set included participants who completed an exit interview and consented to audio recording, as well as participants contributing free-text survey responses.

Participant characteristics were summarized descriptively using counts and percentages for categorical variables and median with interquartile range for continuous variables. The primary analysis used a linear mixed-effects model for overall utility rating, with fixed effects for report type and random intercepts for participant and vignette. Standard OpenEvidence was the reference category. The model was specified as:

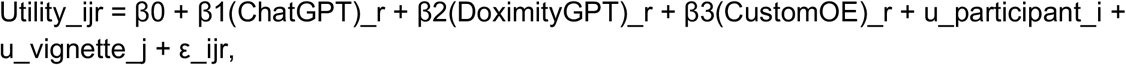

where u_participant_i and u_vignette_j are normally distributed random intercepts for participant and vignette, respectively. The primary test evaluated β3, corresponding to the adjusted mean difference in overall utility between the customized report and standard OpenEvidence. The primary type I error rate was set at α = 0.05 for this single prespecified comparison. Model estimates are reported as adjusted mean differences with 95% confidence intervals and p values.

Secondary report-type contrasts were estimated from the same model. Subcategory ratings were analyzed using analogous models fit separately for each subdomain. Best and worst report selections were summarized as counts and percentages and tested against a uniform 25% choice distribution using chi-squared goodness-of-fit tests, supplemented by exact multinomial confidence intervals when appropriate. Preferred report length was summarized descriptively using counts, percentages, and graphical displays.

A sensitivity analysis additionally adjusted for participant training level. Missing utility ratings were handled using all available observed ratings under a missing-at-random assumption conditional on included model terms. We summarized item-level missingness by report type and vignette.

All analyses were conducted using R version 4.5.3 including the tidyverse set of packages for data manipulation, lme4 and lmerTest for modeling, and gtsummary and ggplot for visualization.

### Qualitative analysis

Qualitative data sources included free-text Qualtrics comments and semistructured exit-interview transcripts. The qualitative analysis used a prespecified prompt-guided workflow to support structured extraction and synthesis. Each interview transcript was processed individually using a transcript-level parsing prompt designed to identify participant-derived meaningful segments, inductive codes, themes, likes, dislikes, product requirements, workflow-integration considerations, and trust and safety observations. The prompt explicitly instructed the model to exclude interviewer statements from thematic analysis, avoid external oncology knowledge, preserve participant anonymity, and output structured JSON.

The individual transcript-level JSON outputs were then combined and analyzed using a prespecified cross-transcript synthesis prompt to identify higher-order themes across participants. Separately, free-text survey comments were pooled by blinded report label and processed using a prespecified sentiment-feature extraction prompt to summarize positive features, negative features, and improvement suggestions for each report subset. The resulting report-specific JSON outputs were then synthesized across report types using a second-order cross-report prompt.

All LLM-assisted qualitative outputs were reviewed by study investigators for interpretability, consistency with source data, and removal of identifying information. Final qualitative findings and design recommendations were derived from the structured JSON outputs after investigator review. The qualitative analysis pipeline utilized GPT 5.4 thinking.

## Data availability

The statistical analysis plan, vignettes, report-generation prompts, qualitative-analysis prompts, and deidentified structured qualitative outputs are available at https://github.com/bbunning/Oncology-LLM-HCI. The pre-specified SAP and prompts are available through OSF: https://osf.io/7qjf4/overview.

## Code Availability

The prompts used for analysis are available at https://osf.io/7qjf4/overview. Additional information can be made available to qualified researchers on reasonable request to the corresponding author.

## Acknowledgements

We are thankful to the Stanford GUIDE team and to members of the HealthRex, ARISE, and Quantitative Sciences Unit for fruitful conversations and insight. This manuscript is partially supported by the Stanford’s Center for Clinical and Translational Education and Research award, under the Biostatistics, Epidemiology and Research Design (BERD) Program: UM1TR004921, by the Clinical and Translational Core of the Stanford Diabetes Research Center: P30DK116074, by the Biostatistics Shared Resource (B-SR) of the NCI-sponsored Stanford Cancer Institute: P30CA124435, The National Library of Medicine (2T15LM007033), and through support of the Gordon and Betty Moore Foundation (Grant #12409). LLMs such as ChatGPT and Claude were utilized in the editing of this manuscript, with review and final approval from the authors.

## Contributions

Conceptualization: BJB, SE, IL, VP, JHC, MD; Supervision: VP, JHC, MD; Funding: JHC, Study Materials: BJB, YW, GH, JEH, MD; Writing: BJB, DJHW, JHC, MD; Experimental Design: BJB, YW, JHC, MD; Data Analysis: BJB, YW, MD; Visualization: BJB, DJHW; Critical Review: BJB, YW, DJHW, GH, JEH, IL, SE, VP, JHC, MD

## Competing Interests

BJB is funded through the National Library of Medicine (2T15LM007033). MD is supported by the Stanford’s Center for Clinical and Translational Education and Research award, under the Biostatistics, Epidemiology and Research Design (BERD) Program: UM1TR004921, by the Biostatistics Shared Resource (B-SR) of the NCI-sponsored Stanford Cancer Institute: P30CA124435, and by the Clinical and Translational Core of the Stanford Diabetes Research Center: P30DK116074.

JHC has received research funding support in part by ARPA-H PACT: Physician-AI Collaboration Teaming, NIH/National Institute of Allergy and Infectious Diseases (1R01AI17812101), NIH-NCATS-Clinical & Translational Science Award (UM1TR004921), Stanford Bio-X Interdisciplinary Initiatives Seed Grants Program (IIP) [R12] [JHC], NIH/Center for Undiagnosed Diseases at Stanford (U01 NS134358), Stanford RAISE Health Seed Grant 2024, Josiah Macy Jr. Foundation (AI in Medical Education), Stanford CARE AI Scholar Fellowship. JHC is Co-founder of Reaction Explorer LLC that develops and licenses organic chemistry education software, is a paid medical expert witness fees via Elite Experts, Paid one-time honoraria or travel expenses for invited presentations by Pfizer, insitro, General Reinsurance Corporation, AASCIF, R1RCM, and other industry conferences, academic institutions, and health systems.

The content is solely the responsibility of the authors and does not necessarily represent the official views of the NIH, Stanford Healthcare, or any other organization.

The remaining authors report no other relevant disclosures.

## Notes

### Competing Interest Statement

BJB is funded through the National Library of Medicine (2T15LM007033).
MD is supported by the Stanford Center for Clinical and Translational Education and Research award, under the Biostatistics, Epidemiology and Research Design (BERD) Program: UM1TR004921, by the Biostatistics Shared Resource (B-SR) of the NCI-sponsored Stanford Cancer Institute: P30CA124435, and by the Clinical and Translational Core of the Stanford Diabetes Research Center: P30DK116074.
JHC has received research funding support in part by ARPA-H PACT: Physician-AI Collaboration Teaming, NIH/National Institute of Allergy and Infectious Diseases (1R01AI17812101), NIH-NCATS-Clinical & Translational Science Award (UM1TR004921), Stanford Bio-X Interdisciplinary Initiatives Seed Grants Program (IIP) [R12] [JHC], NIH/Center for Undiagnosed Diseases at Stanford (U01 NS134358), Stanford RAISE Health Seed Grant 2024, Josiah Macy Jr. Foundation (AI in Medical Education), Stanford CARE AI Scholar Fellowship. JHC is Co-founder of Reaction Explorer LLC that develops and licenses organic chemistry education software, is a paid medical expert witness fees via Elite Experts, Paid one-time honoraria or travel expenses for invited presentations by Pfizer, insitro, General Reinsurance Corporation, AASCIF, R1RCM, and other industry conferences, academic institutions, and health systems.
The content is solely the responsibility of the authors and does not necessarily represent the official views of the NIH, Stanford Healthcare, or any other organization.
The remaining authors report no other relevant disclosures.

### Author Declarations

The Institutional Review Board of Stanford University determined this study to be exempt status human subjects research under protocol 85187.

