## Supplement for "Drivers of Oncologist Preference of AI-Generated Literature Review in a Randomized Mixed-Methods Study"

### Supplemental Results

| | |
|---|---|
| **AI use frequency** | |
| About half the time | 2 (6.1%) |
| Always | 3 (9.1%) |
| Barely | 5 (15%) |
| Consistently as a case warrants it | 6 (18%) |
| Most of the time | 5 (15%) |
| Never | 1 (3.0%) |
| Sometimes | 11 (33%) |
| **AI risk tolerance** | |
| I am incredibly conservative and will wait until more data is available about these tools before using them | 2 (6.1%) |
| I am skeptical but have tried a few clinical use cases for AI | 10 (30%) |
| I use some popular tools consistently | 13 (39%) |
| I am a power user of some AI tools | 4 (12%) |
| I am an early adopter and tinker with all new tools | 4 (12%) |
| **Uses ChatGPT** | 22 (69%) |
| **Uses OpenEvidence** | 27 (84%) |
| **Uses DoximityGPT** | 4 (13%) |
| **Uses UpToDate AI** | 7 (22%) |
| **Uses Claude** | 7 (22%) |
| **Uses Perplexity** | 4 (13%) |
| **Uses NotebookLM** | 3 (9.4%) |

[1] n (%); Mean (SD)

**Table S1 Demographics extended.**

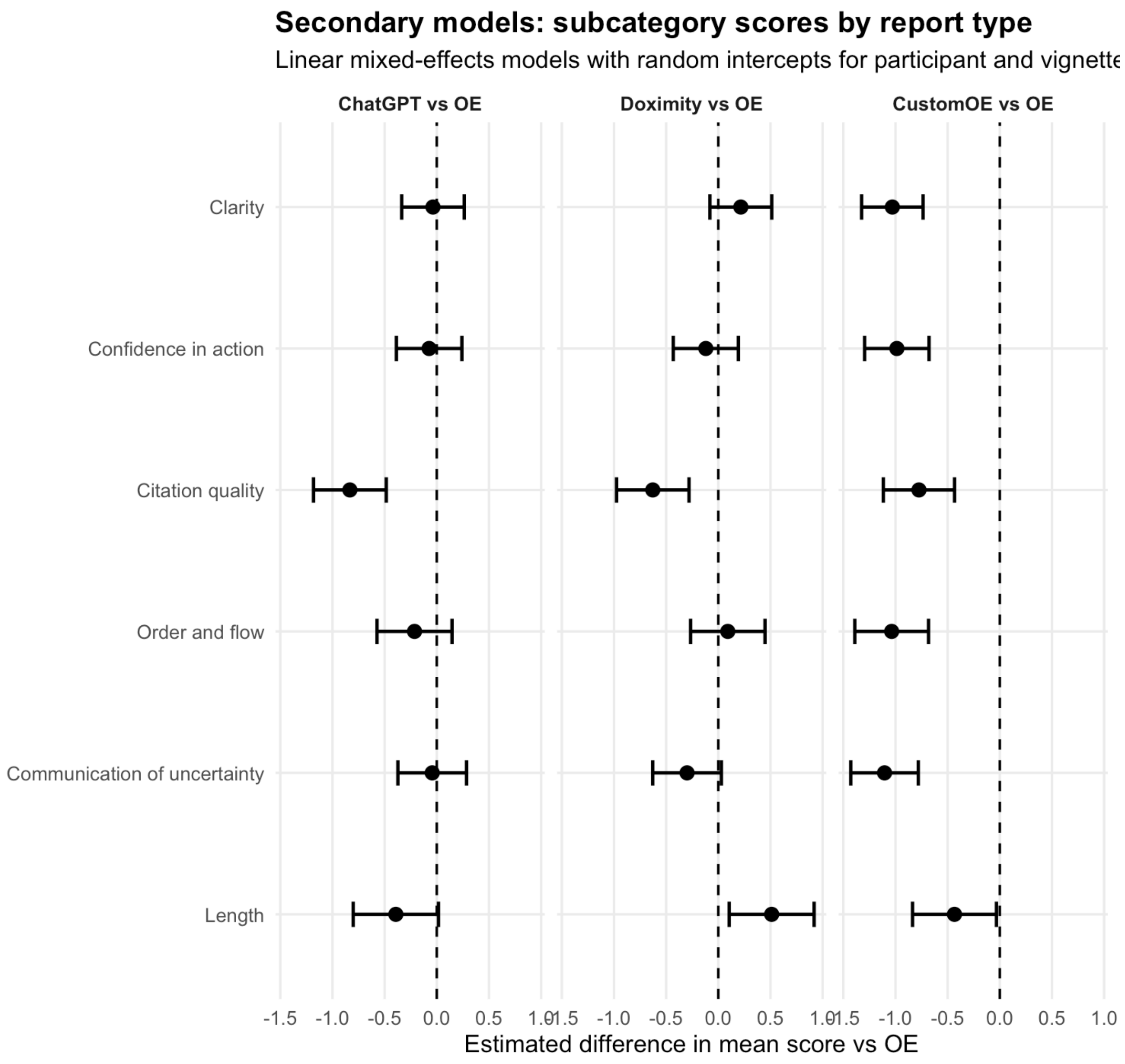

**Figure S1:** Secondary models: pairwise comparisons between models against OpenEvidence against subcategories.

Of note, OpenEvidence has improved perception of citation quality compared to other models studied. Doximity had improved perception of length compared to OpenEvidence. Our custom model performed worse across all subcategories.

```
Structured Cross-Interview Synthesis
│
├── Executive Summary
│   ├── Narrative summary
│   ├── Top-line findings
│   └── Overall interpretation
│
├── Cross-Interview Themes
│   ├── Theme name
│   ├── Theme definition
│   ├── Supporting interview count
│   ├── Representative verbatim quotes
│   ├── Prevalence
│   ├── Importance
│   ├── Direction of sentiment
│   ├── Areas of agreement/disagreement
│   └── Confidence assessment
│
├── Design & Implementation Findings
│   ├── Synthesized likes
│   ├── Synthesized dislikes
│   ├── Design requirements
│   │   ├── Priority
│   │   ├── Rationale
│   │   └── Risk if omitted
│   ├── Workflow synthesis
│   │   ├── Intended use cases
│   │   ├── Workflow constraints
│   │   ├── Format preferences
│   │   └── Current workarounds
│   └── Trust & safety synthesis
│       ├── Trust drivers
│       ├── Trust eroders
│       ├── Verification behaviors
│       └── Medico-legal considerations
│
├── Cross-Case Interpretation
│   ├── Tensions & tradeoffs
│   ├── Minority viewpoints
│   ├── Evidence map
│   └── Publication-ready synthesized statements
│
└── Methodological Summary
    ├── Cross-case interpretation
    ├── Areas of convergence
    ├── Areas of divergence
    └── Study limitations
```

#### Figure S2: Prespecified JSON data structure of Structured Qualitative Knowledge base

A priori defined data structure for transcript interview data. Every synthesized finding above includes: 1) number of supporting interviews for a claim, 2) supporting interview identifiers, 3) representative anonymized verbatim quotes attributed to a claim, 4) traceable references down to the individual interview level data, and 5) a confidence assessment

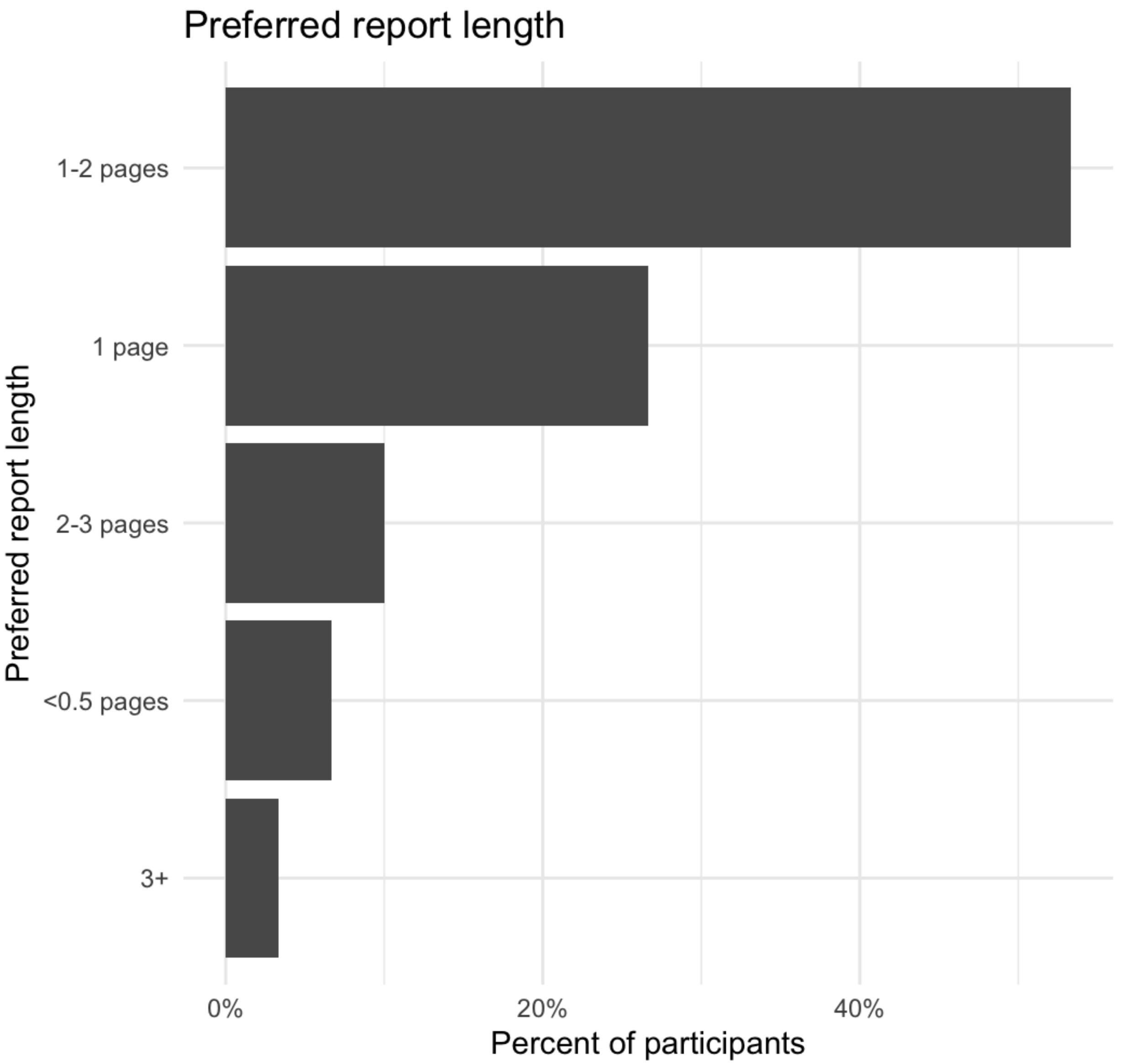

Figure S3: Preferred report length as reported by participants.

#### AI Executive Summary

Verbatim summaries pulled from Knowledge Base JSON:
“Across interviews, participants consistently positioned AI literature-review reports as support tools for speed, orientation, and evidence retrieval rather than as autonomous decision-makers. The most recurrent request was for shorter, more scannable outputs that fit between-patient workflow, often described as a one-page or top-loaded summary with the bottom line first. Clinicians valued reports that made evidence easy to verify, especially when they included recognizable guidelines, visible study type, direct links, and concrete trial outcomes such as PFS, OS, toxicity, and subgroup-relevant details. Trust was strengthened when reports surfaced

high-yield evidence clearly, but weakened quickly by citation mismatch, buried provenance, inaccurate evidence labeling, overconfident tone, or prose that appeared to interpret beyond the source material. Participants repeatedly preferred AI to organize, filter, and contextualize evidence while leaving final clinical judgment with the clinician. Many participants wanted patient-specific framing, but only when grounded in the actual scenario rather than presented as generic or overly decisive recommendations. Formatting mattered substantially: sectioned reports, concise bullets, and selective tables were often welcomed, whereas long paragraphs, crowded reference dumps, and visually noisy tables increased cognitive load. At the same time, there was no single universally preferred format; several participants explicitly wanted a hybrid or customizable output that could adapt to the task, time available, and user familiarity. Less common but important concerns included medico-legal risk, PHI/compliance, the danger of junior users overtrusting polished summaries, and the possibility that AI could confidently answer despite missing critical patient context."

##### AI Overall interpretation

“Taken together, the interviews suggest that oncology clinicians see value in AI literature-review tools when they reduce search burden and improve evidence accessibility without obscuring provenance or overstepping into unsupported decision-making. The most acceptable designs appear to be those that pair concise clinical relevance with transparent, checkable evidence and preserve clinician control over interpretation.”

### Supplemental Qualitative Results

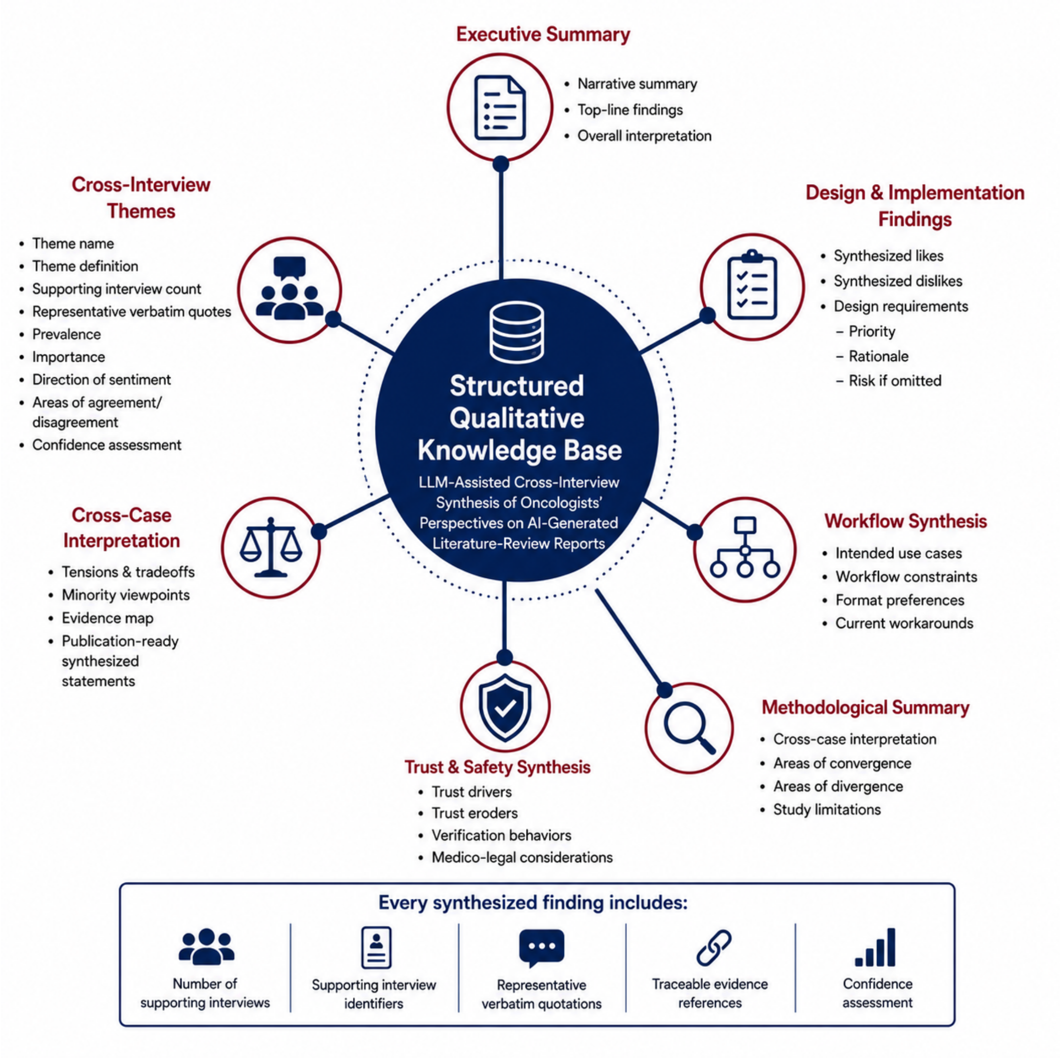

**Figure S4:** Overview of pre-specified categories in the structured knowledge base.

The structured qualitative knowledge base contains the cross-interview themes, synthesized preferences, design requirements, workflow findings, trust-and-safety findings, tensions or tradeoffs, minority perspectives, and representative quotations generated through the prespecified analysis pipeline. The complete deidentified file and prompt set are available through a public repository.

| Domain | Synthesized finding | Supporting interviews, n (%) | Design or governance implication |
|---|---|---|---|
| Verification behavior | Clicking through references or searching for the cited paper directly. | 3 (15%) | Optimize for one-click verification. |
| Verification behavior | Checking AI summaries against NCCN or other guideline sources. | 3 (15%) | Make guideline comparison explicit. |
| Verification behavior | Using personal clinical knowledge, colleagues, or existing workflow references as a backstop. | 3 (15%) | Position output as decision support, not authority. |
| Verification behavior | Inspecting whether the cited study population and outcomes truly match the patient scenario. | 2 (10%) | Surface applicability and patient-match details. |
| Medico-legal/policy concern | Risk that less experienced users may overtrust confident but incomplete or incorrect output. | 2 (10%) | Consider safeguards, training, and visible uncertainty. |
| Medico-legal/policy concern | Need to preserve clinician responsibility for final decisions rather than delegating judgment to AI. | 2 (10%) | Retain clinician-in-the-loop accountability. |
| Medico-legal/policy concern | PHI, HIPAA, and institutional-compliance uncertainty around entering patient information into tools. | 2 (10%) | Clarify privacy, data-use, and institutional compliance. |
| Medico-legal/policy concern | Documentation and insurance justification may require explicit evidence support and clear guideline stance. | 2 (10%) | Provide exportable evidence trails and guideline language. |

#### Table S2. Trust and Safety Synthesis

Interview-level synthesis of features that increased or reduced clinicians' trust in AI-generated literature reports. Trust was supported by recognizable guideline sources, quantitative study outcomes, accurate and accessible citations, and neutral presentation. Trust was undermined by citation mismatch or fabrication, overconfident recommendations, evidence misclassification, and editorialized or overtly AI-generated language. Counts indicate the number of interviews containing supporting evidence for each theme.

##### Selected results from the Structured Qualitative Knowledge Base

Themes:

1. Brevity and scannability are prerequisites for clinical workflow fit
2. Trust depends on visible, high-quality, and clinically interpretable evidence
3. AI is acceptable as support and efficiency help, not as the final decision-maker
4. Usefulness increases when reports are tailored to the patient, task, and user
5. Readable structure matters, but preferred formats differ
6. Trust is fragile when reports are overconfident, inaccurate, or weak on uncertainty

Synthesized likes

1. Participants valued concise reports that deliver the bottom line quickly and minimize fluff.
2. Participants valued visible study-level evidence, especially quantitative outcomes such as PFS, OS, toxicity, and trial phase/type.
3. Participants valued guideline anchoring, evidence hierarchy, and recognizable source names as trust builders.
4. Participants valued sectioned organization that separated efficacy, toxicity, comparisons, and references into an easy-to-follow flow.
5. Participants valued patient-specific or practically framed outputs when they felt grounded in the actual case and useful for next-step thinking.

Synthesized dislikes

1. Participants disliked reports that were too long, repetitive, or paragraph-heavy for real clinical workflow.
2. Participants disliked crowded reference dumps, visually noisy layouts, and tables that added parsing burden instead of clarity.
3. Participants disliked AI-sounding, conversational, or editorialized tone that made the report feel less professional or trustworthy.
4. Participants disliked overconfident or overly directive recommendations that appeared to substitute for clinician judgment.
5. Participants disliked citation mismatch, weak source traceability, and

### Study Materials:

#### Interview Script:

Hi, thank you for taking the time to participate today. My name is Bryan Bunning, and I am a PhD researcher at Stanford University. We are conducting a research study examining how AI-generated literature review reports may support oncology clinicians in reviewing evidence and making clinical decisions. Before we begin, I want to emphasize a few important points:

- This study is **not a test of your medical knowledge, expertise, or decision-making abilities**.
- We are evaluating the reports themselves, not evaluating you.

- There are no right or wrong answers.
- We are interested in your honest reactions and feedback as a practicing oncology clinician.

For the purposes of this study, I'd like you to approach these reports as if you were reviewing them during a busy clinical workday. You are not in a library doing research, so read at a speed that you would in clinic. As you review the reports, we are primarily interested in your **high-level impressions and general feedback**. Please think about questions such as:

- What do you like or dislike about the report structure?
- What information is most useful?
- What information is missing?
- What aspects would make the report more or less useful in your clinical workflow?
- How easy is the report to navigate and understand?
- What specific features did you like or dislike?

We intentionally want zoomed out feedback, looking at this from a "10,000-foot view", rather than asking you about the specifics of the case.

For each clinical scenario, you will review **four different color-coded literature review reports**. The color is consistent between vignettes. I will tell you which report is which at the end.

At the end of the study, I will ask you about the four colored reports, such as:

1. Which report you preferred overall.
2. Which report you liked least.
3. Why you made those selections.

We are particularly interested in understanding the features and design choices that influenced your preferences.

We will ask you for some baseline demographic information and some opinions about AI before we get into the AI generated cases. After you finish the review and survey, we will have an open discussion where I want to hear your thoughts and critiques.

If everything sounds good, we'll begin with the first case.

#### Survey Interview Transcript

(Note: This was interviewee led, with a number of baseline questions and then discussing in depths any points or nuances the interviewee decided to surface)

Thank you for taking the time to go through those different reports. Seeing different cases and a variety of reports provides a foundation for rich discussion and valuable feedback.

Bank of questions to spur discussion:

- To get started, was there anything extreme, in the positive or negative ends, that stood out to you?
- Which color report did you like the best? Why?
- Which color report did you like the least? Why?
- If you could make your own AI report, where you can frankenstein together different pieces of each colored report, what would it look like?
- Do you use AI clinically today? What for? Why or why not?
- Length?

#### Post Survey- Post Interview Transcript:

Thank you again for taking the time to complete this one-hour survey on AI-enabled literature review for clinical practice in oncology. We really appreciate your thoughtful input and the perspective you bring as a practicing oncologist and your feedback will directly inform how these tools should be designed and evaluated for real-world clinical use. For transparency, the reports were blinded using the following color scheme: **Pink** = our custom converted OpenEvidence response, **Red** = ChatGPT response, **Blue** = OpenEvidence response, and **Yellow** = Doximity DoxGPT response.

Do you have any final questions or comments for me?

### Custom Report:

The custom report was created by taking the OpenEvidence response to a vignette question as input, and using a prompt and GPT 5.2 thinking to adapt the prompt.

#### Custom report Prompt:

Reformat the following information into a color-coded, evidence-ranked table, ordered from highest to lowest level of evidence according to the hierarchy below:

Hierarchy (highest → lowest):

Guideline
Clinical Trial Meta-analysis
Phase 3 (P3) Trial
Phase 2 (P2) Trial
Phase 1 (P1) Trial
Large Cohort Study
Cohort Study
Animal Model

Each row should include:
Rank (with color icon)
Level of Evidence
Study Title
Authors
Journal (Year)
Notes

Add a 1–2 sentence summary paragraph with citations, denoted by first author above the table and include a recommendation above the table of citations given the evidence to answer the primary question, guided by the evidence. The title of the table should be “References supporting summary ordered by strength of evidence:” No wording should be below the table. Ensure the evidence hierarchy is enforced!!!
If a level of evidence has no entries, include an empty placeholder row for it.

Template:
(Add a 1–2 sentence summary paragraph here — for example: “This table summarizes all available evidence on [topic], organized from highest to lowest level of rigor. Guideline and meta-analysis evidence are highlighted in green, while preclinical animal data appear in brown. Given this data, include a recommendation given the evidence to answer the primary question, guided by the evidence.”)

References supporting summary ordered by strength of evidence:

| **Rank** | **Level of Evidence** | **Study Title** | **Authors** | **Journal (Year)** | **Notes** |
| --- | --- | --- | --- | --- | --- |
| 🟩 **1** | **Guideline** | | | | |
| 🟢 **2** | **Clinical Trial Meta-analysis** | | | | |
| 🟨🟢 **3** | **Phase 3 (P3) Trial** | | | | |
| 🟨 **4** | **Phase 2 (P2) Trial** | | | | |
| 🟧 **5** | **Phase 1 (P1) Trial** | | | | |
| 🟧🔴 **6** | **Large Cohort Study** | | | | |
| 🟥 **7** | **Cohort Study** | | | | |
| 🟫 **8** | **Animal Model** | | | | |

Context to be reformatted:
QUESTION:
[VERBATIM VIGNETTE GOES HERE]
ANSWER:
[OpenEvidence Response, including all citations, go here]

#### Assessment of custom report against original OpenEvidence Response:

In a post-hoc analysis, we compared the original OpenEvidence responses, specifically the references, to the custom report used in the study. Overall, 77% of references were retained across the vignettes, with ⅗ with 100% retention. No outside references were included in the custom report prompt. The model prompt compressed, dropped, relabeled, and renamed; it did not fabricate sources.

Interestingly, the glioblastoma case dropped a large number of references in the response. The glioblastoma case, discussing the potential efficacy of gabapentin (a drug indicated to address symptom). The specifics of the case, or the references, may be related to its outlier dropping of references..

| **Vignette** | **Original reference count** | **Retained** | **Retitled/renamed** | **Dropped** | **References added from outside sources** |
|---|---|---|---|---|---|
| Prostate | 15 | 15 (100%) | 0 | 0 | 0 |
| Breast | 7 | 7 (100%) | 1 | 0 | 0 |
| Glioblastoma | 16 | 4 (25%) | 1 | 12, including the EANO guideline | 0 |
| GCT | 17 | 15 (88%) | 6 | 2 (both | 0 |

| Vignette | Original reference count | Retained | Retitled/renamed | Dropped | References added from outside sources |
|---|---|---|---|---|---|
| | | | | review articles) | |
| Lymphoma | 5 | 5 (100%) | 3 | 0 | 0 |
| **Total** | **60** | **46 (77%)** | **11** | **14** | **0** |

### Pre-registered LLM prompts:

#### Transcript Parse

You will analyze ONE interview transcript from a mixed-methods study about AI-generated literature review report generation for oncology care workflows.

STUDY GOAL:
Identify themes, likes, dislikes, and actionable improvements for AI literature-review report generation that would increase utility, trust, and fit within oncology clinical workflows.

INDUCTIVE ANALYSIS REQUIREMENT:
Use an inductive approach: derive codes and themes from what the PARTICIPANT (oncologist) says in this transcript. Do NOT impose external frameworks (e.g., TAM/UTAUT) unless the participant explicitly invokes them. Do NOT use prior transcripts, general oncology knowledge, or assumptions.

INTERVIEWER EXCLUSION RULE (CRITICAL):
Bryan Bunning is the interviewer. His comments/questions are NOT data for thematic analysis.

- Exclude Bryan Bunning's statements from coding, themes, likes/dislikes, requirements, and memos.
- You MAY use interviewer prompts only to understand what the participant is responding to, but do not treat them as evidence.
- If speaker labels are missing, treat any question-like, leading, or explanatory text as "interviewer" and exclude it unless the participant repeats/endorses it in their own words.

ANONYMIZATION RULE (CRITICAL):
Preserve participant anonymity in the JSON output.

- Do NOT include any personal names of participants, colleagues, patients, family members, staff, or institutions/locations that could identify a person (e.g., specific hospitals/clinics, unique teams) anywhere in the JSON.
- Do NOT include participant initials, email addresses, phone numbers, usernames, exact addresses, or other identifiers.
- Replace any identifying proper nouns in participant excerpts with bracketed placeholders while keeping meaning:
  - Person → "[NAME]"
  - Patient → "[PATIENT]"
  - Hospital/Clinic/Institution → "[INSTITUTION]"
  - City/State/Country or other location → "[LOCATION]"
  - Unique program/unit/team → "[SERVICE]"
  - Email/phone/ID → "[IDENTIFIER]"
- Tool/product names that are not identifying individuals (e.g., Epic, OpenEvidence, NCCN, SecureGPT) may be kept unless the participant ties them to a uniquely identifying local instance; if so, replace with "[SYSTEM]".
- If a quote cannot be anonymized without losing its meaning, set representative_quote to null and explain briefly in the segment rationale that it was withheld for anonymization (do not add extra JSON keys).
- transcript_id must be anonymized (no names); use a non-identifying ID like "TX001" or a hash-like string you invent.

UNIT OF ANALYSIS:
A "meaningful segment" of participant speech (1–6 sentences) that expresses a single idea or closely related ideas. Use timestamps if available.

IMPORTANT CONSTRAINTS:

- Use ONLY information present in the transcript.

- Do NOT add external oncology knowledge or guidelines not explicitly mentioned by the participant.

- Do NOT invent quotes, speakers, motivations, or clinical facts.

- Quotes must be verbatim participant excerpts (no paraphrasing as quotes) EXCEPT for required anonymization placeholders as specified above.

- If a point is ambiguous, label it “uncertain/ambiguous” and explain why.

- If the transcript does not contain enough evidence for a category, output “Not observed in this transcript.”

INPUT TRANSCRIPT:
Transcript is attached
(If the transcript includes timestamps, preserve them.)

OUTPUT FORMAT (STRICT):
Return JSON ONLY, matching the schema below exactly (no extra keys, no commentary).

JSON SCHEMA:
{
"transcript_id": "[ANONYMIZED ID YOU PROVIDE]",
"interview_context": {
"participant_role": "oncologist|fellow|attending|unknown",
"setting_or_workflow_context": "free text or 'unknown' (must be non-identifying; avoid named places/institutions)",
"tools_mentioned_by_participant": ["list of tools/systems mentioned verbatim by participant, if any (non-identifying tools ok)"]
},
"segments": [
{
"segment_id": "S1",
"timestamp_start": "if available else null",
"timestamp_end": "if available else null",
"speaker": "participant",
"verbatim_excerpt": "exact participant text from transcript (<=60 words), anonymized with placeholders if needed",
"codes": ["code_1", "code_2"],
"valence": "positive|negative|mixed|neutral",
"object_of_evaluation": "e.g., length, tone, evidence display, tables, citations, workflow fit, trust, UI, visuals, guideline content, etc.",
"rationale": "1–2 sentences explaining why you assigned these codes using ONLY the excerpt (may note anonymization if quote withheld)."
}

],
"codebook": [
{
"code": "string_snake_case",
"definition": "operational definition grounded in participant language (1–2 sentences)",
"inclusion_criteria": ["bullets"],
"exclusion_criteria": ["bullets"],
"example_segment_ids": ["S1","S7"]
}
],
"themes": [
{
"theme_id": "T1",
"theme_name": "short descriptive name using participant-near language when possible",
"theme_definition": "2–4 sentences grounded in participant statements (no outside concepts)",
"supporting_segments": ["S1","S3","S9"],
"representative_quotes": [
{
"segment_id": "S1",
"quote": "verbatim participant quote (<=35 words), anonymized with placeholders if needed, or null if cannot be safely anonymized"
}
],
"overall_valence": "positive|negative|mixed|neutral",
"confidence": "high|medium|low",
"confidence_basis": "e.g., repeated across multiple participant segments vs single mention"
}
],
"likes": [
{
"like_id": "L1",
"statement": "concise statement of what the participant liked",
"supporting_segments": ["S2","S8"],
"representative_quote": "verbatim participant quote (<=25 words), anonymized with placeholders if needed, or null if cannot be safely anonymized"
}
],
"dislikes": [
{
"dislike_id": "D1",
"statement": "concise statement of what the participant disliked",
"supporting_segments": ["S4"],
"representative_quote": "verbatim participant quote (<=25 words), anonymized with placeholders if needed, or null if cannot be safely anonymized"

```
}
],
"requirements": [
{
"req_id": "R1",
"type": "must_have|nice_to_have|avoid",
"requirement": "single sentence requirement phrased as a product/design constraint derived from participant statements",
"supporting_segments": ["S4","S10"],
"risk_if_ignored": "1 sentence"
}
],
"workflow_integration": {
"current_workarounds": ["participant-described alternatives/workarounds, if any"],
"intended_use_cases": ["participant-described use cases, if any"],
"constraints": ["participant-mentioned constraints: time, cognitive load, trust, documentation, etc."],
"output_format_preferences": ["participant-mentioned formatting preferences"]
},
"trust_and_safety": {
"trust_drivers": ["participant-mentioned trust builders"],
"trust_erosions": ["participant-mentioned trust reducers"],
"verification_behaviors": ["participant-described checking behaviors, if mentioned"],
"medico_legal_or_policy_concerns": ["participant-mentioned concerns, else empty"]
},
"quantified_mentions": {
"top_codes_by_count": [
{"code": "string_snake_case", "count": 0}
],
"notes": "Counts are approximate; note any ambiguity due to transcript format and anonymization replacements."
},
"analytic_memos": {
"summary": "5–8 sentences summarizing the participant perspective only (must be non-identifying; no names/places).",
"tensions_or_tradeoffs": ["tradeoffs explicitly or implicitly described by the participant"],
"open_questions_for_next_interview": ["2–5 probes based on what this participant raised"]
}
}
```

CODING GUIDANCE:

- Only code PARTICIPANT speech.
- Use inductive, descriptive codes that stay close to participant wording.

- Create new codes when new ideas emerge; reuse existing codes within the transcript when the same idea recurs.

- Keep the codebook typically 10–25 codes per transcript.

Now produce the JSON.

#### Multi-Transcript Synthesis Prompt:

You will analyze a SET of JSON summaries generated from individual interview transcripts in a mixed-methods study about AI-generated literature review report generation for oncology care workflows.

### STUDY GOAL

Synthesize findings across multiple oncologist interviews to identify cross-participant themes, recurring likes and dislikes, workflow needs, trust considerations, and design implications for AI-generated literature review reports used in oncology care.

### ANALYTIC TASK

You are performing a **cross-case qualitative synthesis** using the structured JSON outputs from individual interviews as the sole input data source.

Your task is to:

- Compare patterns across participants
- Aggregate overlapping codes, likes, dislikes, requirements, workflow constraints, and trust/safety observations
- Identify higher-order synthesized themes grounded in the interview JSONs
- Produce an executive summary and publication-ready synthesized statements
- Preserve nuance, including disagreement, minority viewpoints, and tensions/tradeoffs

### INDUCTIVE SYNTHESIS REQUIREMENT

Use an inductive approach. Build synthesis from the content of the provided interview JSONs only.

- Do NOT introduce external frameworks (e.g., TAM, UTAUT, human factors models) unless they are explicitly present in the interview outputs.
- Do NOT add outside oncology knowledge, product design assumptions, or clinical workflow assumptions not reflected in the input JSONs.
- Do NOT infer participant intent beyond what is reasonably supported by the source JSONs.
- Do NOT collapse distinct ideas into one theme if meaningful differences remain.

### INPUT DATA RULES

The input consists of multiple interview-summary JSON objects, each derived from one participant interview.
Assume each JSON may contain:

- segments
- codebook
- themes
- likes
- dislikes

- requirements
- workflow_integration
- trust_and_safety
- quantified_mentions
- analytic_memos

Use these interview JSONs as the analytic record.

### INTERVIEWER EXCLUSION RULE

Bryan Bunning is the interviewer and is not part of the analyzed participant data.
If any interviewer-originated ideas appear to have been improperly carried into an interview JSON, do not elevate them unless clearly endorsed by participant-derived evidence in that JSON.

### UNIT OF SYNTHESIS

The primary unit of synthesis is the **participant-level interview JSON**.
The secondary units are:

- coded segments
- interview-level themes
- likes/dislikes
- requirements
- workflow/trust observations

### IMPORTANT CONSTRAINTS

- Use ONLY information present in the provided interview JSONs.
- Do NOT invent quotations.
- Do NOT create false consensus.
- When participants differ, explicitly describe the divergence.
- Preserve minority but salient viewpoints if they have clear implications.
- Distinguish:
  - commonly mentioned findings
  - strongly expressed findings
  - design-critical findings
- If support is weak or inconsistent, label it accordingly.
- If a category is not sufficiently supported across interviews, output: "Insufficient cross-interview evidence."

### SYNTHESIS GUIDANCE

When synthesizing:

- Merge semantically overlapping codes into broader synthesized concepts only when justified by the source material.
- Preserve original participant-near language when possible.

- Note the number of interviews contributing to a theme.
- Treat frequency as informative but not definitive; a less frequent issue may still be important if it is concrete, high-risk, or strongly stated.
- Pay special attention to:
    - workflow fit
    - time burden
    - trust and verification
    - evidence display and citations
    - report organization
    - visual formatting
    - clinical usefulness
    - cognitive load
    - ambiguity tolerance
    - medico-legal concerns
    - perceived risks of misleading output
    - desired safeguards and product requirements

### INPUT

[PASTE ARRAY OF INDIVIDUAL INTERVIEW JSON OBJECTS HERE]

### OUTPUT FORMAT

Return JSON ONLY, matching the schema below exactly.

{
"synthesis_metadata": {
"study_focus": "AI-generated literature review report generation for oncology care workflows",
"num_interviews_synthesized": 0,
"num_interviews_with_usable_data": 0,
"notes_on_input_quality": "Brief note on missingness, inconsistent detail, or other limitations across interview JSONs."
},
"executive_summary": {
"summary_paragraph": "6-10 sentence publication-ready summary synthesizing the overall findings across interviews.",
"top_line_findings": [
"3-7 concise high-level findings suitable for a results section or abstract"
],
"overall_interpretation": "2-4 sentences interpreting what these findings suggest about the design and implementation of AI literature review tools in oncology workflows, grounded only in the input data."
},
"cross_interview_themes": [
{
"synth_theme_id": "ST1",
"theme_name": "short descriptive name",

```
"theme_definition": "3-5 sentences describing the synthesized theme across interviews",
"contributing_interviews": ["transcript_id_1", "transcript_id_2"],
"num_interviews_supporting": 0,
"supporting_evidence": {
"supporting_theme_ids": ["T1", "T2"],
"supporting_segment_ids": ["transcript1:S3", "transcript4:S8"],
"supporting_like_ids": ["transcript2:L1"],
"supporting_dislike_ids": ["transcript3:D2"],
"supporting_requirement_ids": ["transcript5:R1"]
},
"representative_quotes": [
{
"transcript_id": "string",
"segment_id": "S1",
"quote": "verbatim quote <=35 words"
}
],
"direction_of_sentiment": "positive|negative|mixed|neutral",
"prevalence": "high|moderate|low",
"importance": "high|moderate|low",
"rationale_for_importance": "Explain whether this theme matters because of recurrence, strength of language, workflow consequences, trust implications, or design risk.",
"variation_or_disagreement": "Describe important differences across participants, or state 'Minimal disagreement observed.'",
"confidence": "high|medium|low",
"confidence_basis": "Explain confidence based on consistency, richness, and number of supporting interviews."
}
],
"synthesized_likes": [
{
"s_like_id": "SL1",
"statement": "Cross-interview synthesized statement of what participants valued",
"contributing_interviews": ["transcript_id_1", "transcript_id_3"],
"num_interviews_supporting": 0,
"example_quotes": [
{
"transcript_id": "string",
"segment_id": "S2",
"quote": "verbatim quote <=25 words"
}
]
}
],
```

"synthesized_dislikes": [
{
"s_dislike_id": "SD1",
"statement": "Cross-interview synthesized statement of what participants disliked",
"contributing_interviews": ["transcript_id_2", "transcript_id_4"],
"num_interviews_supporting": 0,
"example_quotes": [
{
"transcript_id": "string",
"segment_id": "S5",
"quote": "verbatim quote <=25 words"
}
]
}
],
"design_requirements": [
{
"design_req_id": "DR1",
"priority": "must_have|should_have|nice_to_have|avoid",
"requirement": "Single sentence product/design requirement synthesized across interviews",
"why_it_matters": "1-2 sentences grounded in participant evidence",
"contributing_interviews": ["transcript_id_1", "transcript_id_2"],
"num_interviews_supporting": 0,
"risk_if_ignored": "1 sentence",
"supporting_evidence": ["transcript1:R2", "transcript3:D1", "transcript4:S7"]
}
],
"workflow_synthesis": {
"intended_use_cases": [
{
"use_case": "synthesized use case",
"contributing_interviews": ["transcript_id_1"],
"num_interviews_supporting": 0
}
],
"workflow_constraints": [
{
"constraint": "synthesized workflow constraint",
"contributing_interviews": ["transcript_id_2"],
"num_interviews_supporting": 0
}
],
"format_preferences": [
{

"preference": "synthesized output-format preference",
"contributing_interviews": ["transcript_id_3"],
"num_interviews_supporting": 0
}
],
"current_workarounds": [
{
"workaround": "participant-described workaround synthesized across interviews",
"contributing_interviews": ["transcript_id_4"],
"num_interviews_supporting": 0
}
]
},
"trust_and_safety_synthesis": {
"trust_drivers": [
{
"driver": "synthesized trust builder",
"contributing_interviews": ["transcript_id_1", "transcript_id_5"],
"num_interviews_supporting": 0
}
],
"trust_erosions": [
{
"erosion": "synthesized trust reducer",
"contributing_interviews": ["transcript_id_2"],
"num_interviews_supporting": 0
}
],
"verification_behaviors": [
{
"behavior": "how participants described checking or validating outputs",
"contributing_interviews": ["transcript_id_3"],
"num_interviews_supporting": 0
}
],
"medico_legal_or_policy_concerns": [
{
"concern": "synthesized concern",
"contributing_interviews": ["transcript_id_4"],
"num_interviews_supporting": 0
}
]
},
"tensions_and_tradeoffs": [

{
"tradeoff_id": "TT1",
"tradeoff": "Concise statement of a tension observed across interviews",
"side_a": "first side of the tension",
"side_b": "second side of the tension",
"contributing_interviews": ["transcript_id_1", "transcript_id_2"],
"interpretation": "1-3 sentences explaining the tension in participant-grounded terms"
}
],
"publication_ready_synthesized_statements": [
{
"statement_id": "P1",
"type": "results|discussion|implication",
"statement": "A polished, publication-ready synthesized statement grounded in the data and suitable for a manuscript.",
"support_level": "strong|moderate|preliminary",
"basis": "Brief explanation of support based on breadth/consistency/richness of interview evidence."
}
],
"minority_or_divergent_views": [
{
"view_id": "MV1",
"statement": "Important but less common or divergent viewpoint",
"contributing_interviews": ["transcript_id_6"],
"why_it_should_be_retained": "Why this viewpoint remains analytically important despite lower prevalence"
}
],
"evidence_map": {
"most_salient_synthesized_codes_or_concepts": [
{
"concept": "string",
"approx_num_interviews": 0,
"related_interview_codes": ["code_a", "code_b"]
}
],
"notes": "Counts are approximate and based on interview-level JSON outputs rather than re-coding full transcripts."
},
"methodological_memo": {
"cross_case_summary": "5-8 sentences summarizing the analytic pattern across participants.",
"areas_of_strong_convergence": [
"list"
],

"areas_of_partial_convergence": [
"list"
],
"areas_of_disagreement_or_heterogeneity": [
"list"
],
"limitations_of_this_synthesis": [
"Important limitations caused by input quality, missing detail, uneven coding depth, or ambiguity."
]
}
}

#### Sentiment Features Prompt:

You will analyze a concatenated set of qualitative comments from practicing oncologists about AI-generated reports and literature-review outputs used in oncology workflows.

### STUDY PURPOSE

The purpose of this analysis is to identify the **strengths, weaknesses, and improvement opportunities** described by the cohort regarding AI-generated reports and literature-review tools for oncology practice.

### UNIT OF ANALYSIS

The input consists of **multiple short free-text comments concatenated together** from one analytic subset, such as:

- one **report type** within a vignette,
- one **report type across multiple vignettes**, or
- **study closeout comments** aggregated across participants.

Treat the full input as a **pooled corpus of comments from multiple oncologists**, not as a single coherent narrative from one speaker.

### ANALYTIC TASK

Extract and synthesize the **positive features**, **negative features**, and **suggested improvements** mentioned in the text.

Your goal is to produce a structured summary of:

1. what oncologists liked,
2. what oncologists disliked,
3. what changes they wanted,
4. and which issues appeared most salient in this subset.

### INDUCTIVE ANALYSIS REQUIREMENT

Use an **inductive qualitative approach**:

- Derive categories from the language and content of the comments themselves.
- Do **not** impose external frameworks unless explicitly supported by the text.
- Do **not** infer attitudes that are not grounded in the input.
- Stay close to the wording and meaning used by the participants.

### IMPORTANT INTERPRETIVE RULES

- These comments may be fragmented, telegraphic, redundant, or inconsistently phrased.
- Normalize minor spelling/grammar issues mentally, but preserve the intended meaning.

- Multiple comments may express the same idea using different wording; consolidate these into a shared feature when appropriate.
- Distinguish between:
  - **positive features** (what worked well),
  - **negative features** (what did not work well),
  - **improvement requests** (what should be changed),
  - and **mixed/conditional sentiments** (e.g., “good content but too long”).
- When a comment contains both praise and criticism, capture both separately.
- Do not over-aggregate distinct concepts into one broad bucket if the text supports finer distinctions.

### REPORT-TYPE AWARENESS

The input belongs to a specific analytic subset defined by the researcher, usually a **report type**.
You should summarize sentiment **within this subset only** and not compare to other report types unless the input explicitly includes such comparisons.

If the input is from study closeout questions rather than one report type, apply the same logic and summarize the pooled sentiment across the closeout corpus.

### OUTPUT REQUIREMENTS

Return valid JSON only.

Use the following schema exactly:

```
{
   "analysis_unit": {
      "subset_type": "report_type | closeout | other",
      "subset_label": "string",
      "notes": "brief description of what this corpus represents"
   },
   "summary": {
      "overall_impression": "2-4 sentence synthesis of the major strengths
and weaknesses in this subset",
      "dominant_positive_themes": ["theme 1", "theme 2", "theme 3"],
      "dominant_negative_themes": ["theme 1", "theme 2", "theme 3"],
     "dominant_improvement_requests": ["request 1", "request 2", "request 3"]
   },
   "positive_features": [
      {
      "feature_label": "concise name of positive feature",
      "description": "brief explanation grounded in the comments",
       "representative_examples": [
      "short paraphrase or brief quote fragment 1",
      "short paraphrase or brief quote fragment 2"
      ],
      "relative_salience": "high | medium | low"
      }
```

```
  ],
  "negative_features": [
    {
    "feature_label": "concise name of negative feature",
    "description": "brief explanation grounded in the comments",
     "representative_examples": [
    "short paraphrase or brief quote fragment 1",
    "short paraphrase or brief quote fragment 2"
    ],
    "relative_salience": "high | medium | low"
    }
  ],
  "improvement_requests": [
    {
    "request_label": "concise name of requested improvement",
    "description": "brief explanation of what participants wanted changed",
     "representative_examples": [
    "short paraphrase or brief quote fragment 1",
    "short paraphrase or brief quote fragment 2"
    ],
    "targeted_problem": "what weakness this request appears to address",
    "relative_salience": "high | medium | low"
    }
  ],
  "mixed_or_tradeoff_comments": [
    {
    "theme_label": "concise name of tension or tradeoff",
    "description": "brief explanation of the mixed sentiment",
     "representative_examples": [
    "short paraphrase or brief quote fragment 1"
    ]
    }
  ],
  "analytic_memos": {
    "interpretive_notes": [
    "brief memo 1",
    "brief memo 2"
    ],
    "data_quality_notes": [
    "note on fragmentation/redundancy/ambiguity if relevant"
    ]
  }
}
```

### CODING GUIDELINES

When grouping comments, favor categories such as:

- clarity
- organization

- brevity vs verbosity
- explicit reasoning
- usefulness of citations
- citation specificity
- transparency of evidence use
- actionability for clinical use
- completeness
- trustworthiness
- workflow fit
- readability
- formatting/structure

Only use these categories if they are supported by the text. You may create other categories when the comments indicate a different concept.

### QUOTATION / EXCERPT RULES

- Use only short excerpts or paraphrased fragments in `representative_examples`.
- Do not fabricate quotes.
- If wording is too fragmentary for a quote, paraphrase faithfully.
- Keep examples concise.

### SALIENCE RULES

Estimate `relative_salience` based on:

- repetition across comments,
- clarity/directness of the sentiment,
- and emphasis in the language.

Do not treat salience as a numeric frequency count unless the data clearly supports that.

### DO NOT

- Do not invent participant intent.
- Do not claim statistical prevalence.
- Do not compare subsets unless explicit in the input.
- Do not introduce external oncology knowledge.
- Do not overinterpret isolated fragments.
- Do not output anything except valid JSON.

#### Multi-Sentiment Synthesis Prompt

You will analyze **multiple JSON outputs** generated by the `sentiment_features.docs` prompt from a mixed-methods study of practicing oncologists evaluating AI-generated reports and literature-review outputs for oncology workflows.

### STUDY PURPOSE

The purpose of this analysis is to synthesize, across report types or study sections, the major **strengths**, **weaknesses**, **requested improvements**, and **cross-cutting tradeoffs** that oncologists described regarding AI-generated reports and literature-review tools.

### INPUT

The input consists of multiple structured JSON objects.
Each JSON object summarizes one analytic subset, such as:

- one **report type** within a vignette,
- one **report type across multiple vignettes**, or
- one **aggregated study closeout corpus**.

Each JSON contains:

- an `analysis_unit`,
- a synthesized `summary`,
- `positive_features`,
- `negative_features`,
- `improvement_requests`,
- `mixed_or_tradeoff_comments`,
- and `analytic_memos`.

### UNIT OF ANALYSIS

Your job is to synthesize **across these subset-level summaries**.

This is a **second-order qualitative synthesis**:

- You are synthesizing across previously extracted themes/features.
- Do **not** return to line-by-line coding of raw participant comments.
- Work only from the provided JSON inputs.

### ANALYTIC TASK

Produce a structured cross-report synthesis that identifies:

1. the most recurring positive features across subsets,
2. the most recurring negative features across subsets,
3. the most recurring improvement requests,

4. the most important tradeoffs/tensions,
5. points that appear common across report types,
6. and points that appear more subset-specific.

### INDUCTIVE SYNTHESIS REQUIREMENT

Use an **inductive, evidence-grounded synthesis approach**:

- Build higher-order categories from the JSON inputs.
- Preserve distinctions when different subsets highlight meaningfully different issues.
- Consolidate overlapping labels when they clearly refer to the same underlying concept.
- Stay close to the content of the source JSONs.
- Do not introduce external frameworks unless explicitly justified by the input content.

### IMPORTANT INTERPRETIVE RULES

- Treat salience as **qualitative recurrence and emphasis**, not as formal quantitative frequency.
- Do not make statistical claims such as "most oncologists" unless explicitly supported by the input.
- Look for both:
  - **cross-cutting themes** that recur across multiple subsets, and
  - **subset-specific themes** that appear more localized.
- When a theme appears in both positive and negative form across subsets, capture the tension explicitly.
  - Example: "comprehensive" may be valued by some comments but criticized as "too long" in others.
- Preserve nuance: do not flatten all themes into generic buckets if the subset-level JSONs contain clinically meaningful distinctions.

### OUTPUT REQUIREMENTS

Return valid JSON only.

Use the following schema exactly:

```
{
   "corpus_summary": {
      "number_of_input_jsons": 0,
      "subset_labels": ["string"],
      "overall_synthesis": "4-6 sentence publication-ready synthesis
describing the major strengths, weaknesses, and design implications across
report types or study sections"
   },
   "cross_cutting_positive_features": [
      {
      "theme_label": "concise name of cross-cutting strength",
      "description": "brief synthesis of how this strength appeared across
subsets",
      "appears_in_subsets": ["subset label 1", "subset label 2"],
```

```
      "qualitative_salience": "high | medium | low",
      "notes_on_variation": "optional note on how this strength differed by subset"
      }
   ],
   "cross_cutting_negative_features": [
      {
      "theme_label": "concise name of cross-cutting weakness",
      "description": "brief synthesis of how this weakness appeared across subsets",
      "appears_in_subsets": ["subset label 1", "subset label 2"],
      "qualitative_salience": "high | medium | low",
      "notes_on_variation": "optional note on how this weakness differed by subset"
      }
   ],
   "cross_cutting_improvement_requests": [
      {
      "theme_label": "concise name of recurring requested improvement",
      "description": "brief synthesis of what participants wanted changed across subsets",
      "appears_in_subsets": ["subset label 1", "subset label 2"],
      "targeted_problem": "what broader weakness this request seems intended to solve",
      "qualitative_salience": "high | medium | low"
      }
   ],
   "key_tradeoffs": [
      {
      "tradeoff_label": "concise name of recurring tension",
      "description": "brief explanation of the tradeoff across subsets",
      "positive_pole": "what users valued",
      "negative_pole": "what users found problematic",
      "appears_in_subsets": ["subset label 1", "subset label 2"]
      }
   ],
   "subset_specific_findings": [
      {
      "subset_label": "string",
       "distinctive_positive_features": ["feature 1", "feature 2"],
       "distinctive_negative_features": ["feature 1", "feature 2"],
       "distinctive_improvement_requests": ["request 1", "request 2"],
      "interpretive_note": "what seemed distinctive about this subset relative to the others"
      }
   ],
   "publication_ready_synthesis": {
      "results_paragraph": "1 paragraph suitable for a mixed-methods Results section describing cross-report feature patterns",
```

```
    "design_implications_paragraph": "1 paragraph suitable for Discussion or
Implications describing what the findings suggest AI report tools should
improve"
  },
  "analytic_memos": {
    "theme_merging_decisions": [
    "brief note explaining important theme consolidations"
    ],
   "uncertainties_or_limitations": [
    "brief note about ambiguity, inconsistent labeling, or limited
comparability across subsets"
    ]
  }
 }
```

### SYNTHESIS GUIDELINES

When merging themes across subset-level JSONs:

- Merge labels that clearly reflect the same idea even if worded differently.
  - Example: "too long," "verbose," and "overly detailed" may be merged into a broader theme such as `excessive length/verbosity`.
- Keep distinct themes separate when they represent different concepts.
  - Example: `lack of citation specificity` should remain distinct from `lack of overall trustworthiness` unless the input clearly combines them.
- Favor labels that are:
  - concise,
  - publication-friendly,
  - and faithful to participant meaning.

### PRIORITY DOMAINS TO WATCH FOR

Only use these when supported by the input:

- organization / structure
- length / concision
- explicit reasoning
- transparency
- citation specificity
- evidence traceability
- readability
- actionability
- completeness
- trustworthiness
- workflow fit
- interpretability
- clinical usefulness

### RESULTS PARAGRAPH STYLE

The `results_paragraph` should read like a manuscript Results section:

- neutral and concise,
- theme-focused,
- no exaggerated claims,
- no unsupported quantification,
- no bullet formatting,
- appropriate for publication in a mixed-methods study.

### DESIGN IMPLICATIONS PARAGRAPH STYLE

The `design_implications_paragraph` should translate the findings into practical implications for AI report design. It should focus on features that would likely improve utility for oncology workflows, based only on the input JSONs.

### DO NOT

- Do not invent findings absent from the subset JSONs.
- Do not return to raw-data style quoting unless brief examples are already embedded in the source JSON logic.
- Do not make numerical prevalence claims unless explicitly supported by the inputs.
- Do not compare subsets beyond what the JSON evidence supports.
- Do not output anything except valid JSON.

ADDITIONAL PUBLICATION ORIENTATION:
- Prefer theme labels that would be understandable in a manuscript table or Results section.
- Emphasize recurring design priorities that cut across report types.
- Surface tensions that are especially relevant to clinical AI adoption, such as comprehensiveness versus brevity, or evidence synthesis versus citation traceability.
- Keep the tone analytical and restrained.

### Statistical Analysis Plan

**Statistical Analysis Plan (SAP)**

***Please complete all relevant sections. Instructions are in red and should be deleted when completing the SAP. The purpose of this template is to provide a general layout, but sections can be reformatted as you wish.***
***The creation of this template was made possible by the Duke CTSA BERD.***

Title: **A randomized vignette study of AI literature review for Oncology**

CRU/Department/Division/Center: Quantitative Sciences Unit, Division of Computational Medicine, Department of Medicine, Stanford

IRB Number: 85187

Primary Investigator: Manisha Desai

Collaborative Lead: Bryan Bunning

Co-authors (if know):
Stanford affiliated: Bryan Bunning, Jessica Hope, Isabel Weng, David J Wu, Vijay Panduragan, Jonathan H Chen, Manisha Desai
UCLA: Gavin Hui

Analysis Biostatistician(s): Bryan Bunning

Subject Matter Expert:
Oncology: Gavin Hui, David J Wu,
Biostatistics: Bryan Bunning, Manisha Desai,
AI: Bryan Bunning, Jonathan H Chen, Vijay Pandurangan

Original Creation Date: March 5 2026

Version Date: April 15 2026

Project Goal(s): Manuscript to informatics aligned journal, generation of feedback to improve AI response utility for oncologists in medical decision making.

Submission Deadline(s): Spring 2026

Investigator Agreement

- ☐ All statistical analyses included in an abstract or manuscript should reflect the work of the biostatistician(s) listed on this SAP. No changes or additional analyses should be made to the results or findings without discussing with the project biostatistician(s).
- ☐ All biostatisticians on this SAP should be given sufficient time to review the full presentation, abstract, manuscript, or grant and be included as co-authors on any abstract or manuscript resulting from the analyses.
- ☐ If substantial additional analysis is necessary or the aims of the project change, a new SAP will need to be developed.
- ☐ If you engaged the CTSA's BERD please ensure you cite UL1TR003142 award when disseminating work. If your study was cancer-related, please ensure you cite P30CA124435 award when disseminating work. If your study was diabetes-related, please ensure you cite P30DK116074 award when disseminating work. These publications should also be submitted to PubMed Central.

☐ I have reviewed the SAP and understand that any changes must be documented.

Acknowledged by:

Lead Principal Investigator: Lead Biostatistician:

_______________________ ______________________

Date: ____________________ Date: ___________________

**Activity Log:**
V0.5 – March 5 2026 Endpoints listed on ORF
V1.0 – March 20 2026- SAP in QSU recommended format
V2.0 – April 15 2026 - improved readability and structure, addition of introduction, switching subcategory assessment to a GLM after internal discussion pre-unblinding.
V2.1 – April 28 2026 – readability (deleting placeholders, font, aesthetics).

#### 1. Study Overview

Background/Introduction:

This exempt human subjects research study evaluates clinician perceptions of multiple generative-AI–produced oncology literature review outputs across multiple oncology vignettes, with the goal of understanding relative utility, preferences, and improvement opportunities. Participants complete a remote session in which they review vignette-specific AI responses from four sources—standard OpenEvidence, a customized prompt/report variant, ChatGPT, and DoximityGPT —and then provide structured ratings and comparative judgments; the study also includes exit interviews recorded with permission. Quantitative analysis will occur mainly through 7 point likert scales collected during the survey. The main qualitative analysis will be through a pre-specified custom prompted LLM that ingests the zoom transcripts and inductively pulls out key themes discussed during participant interviews. The LLMs are a powerful tool that can take unstructured interviews into structured data for parsing and assessment.

The study's stated purpose is to compare and contrast AI tools available to clinicians and to learn clinician preferences and perceived utility in oncology workflows, with the intent of informing better integration and design of the next generation of AI-assisted literature review.

#### Introduction:

Oncologists practice in an environment where clinical evidence expands rapidly, guidelines evolve frequently, and the set of available treatment options continues to grow in complexity. A longitudinal analysis of oncology guideline content has shown substantial increases in the length and complexity of major oncology clinical practice guidelines over time, reflecting the widening breadth of decision-relevant information that clinicians must interpret and apply. (1) [1] In parallel, the operational demands of modern cancer care, including a growing electronic health record (EHR) communication burden, constrain the time available for evidence review and deep appraisal. National longitudinal data demonstrate increasing inbox message volume and EHR workload across oncology specialties from 2019 to 2022, with medical oncology/hematology among the most burdened groups. (2) [2] These realities align with broader research on point-of-care learning: clinicians frequently cite insufficient time, the volume of available information, uncertainty about which resources to search, and the perceived effort of searching as barriers to answering clinical questions during care. (3,4) [20]

Large language models (LLMs) have recently emerged as a new class of retrieval-and-synthesis tools that can generate natural-language summaries, propose reasoning chains, and draft text in response to open-ended prompts. Their rapid diffusion into healthcare has been driven by the promise of reducing knowledge-search time, supporting evidence synthesis, and lowering the friction of documentation and communication tasks. Yet early evaluations indicate that real-world value is context-dependent. For example, in a retrospective primary-care evaluation, OpenEvidence generated evidence-based recommendations that aligned with physician plans and received high ratings for clarity, relevance, and evidence support, while the study emphasized the need for prospective evaluation, more complex cases, and multidisciplinary settings to understand clinical impact. (5) [5] Surveys in radiation oncology similarly suggest active, role-dependent adoption of ChatGPT for administrative and writing-related tasks alongside persistent concerns about accuracy, privacy, and ethical use in cancer settings. (6) [6] Collectively, these findings suggest that clinicians are already experimenting with LLMs, but that the field lacks detailed, workflow-grounded understanding of *how* clinicians integrate these systems into care, *which workflows* are prioritized, and *where friction points* and safety risks concentrate particularly in oncology, where evidence synthesis is both frequent and high-stakes.

A key translational challenge is that LLM efficacy in medicine is not solely a function of model capability; it is also shaped by interaction design, cognitive biases, and the structure of human–AI collaboration. Decades of human-factors research show that automation changes attention allocation and decision processes, producing risks of inappropriate reliance. The conceptual framework of automation “use, misuse, disuse, and abuse” highlights that overreliance can lead to monitoring failures and biased decision-making, while underuse can prevent potential gains. (7) [7] Empirically, automation bias has been demonstrated through omission errors and commission errors, including scenarios where users follow imperfect automated recommendations even when conflicting, valid indicators are available. (8) [8] In clinical decision support, systematic evidence indicates that new error modes can be introduced even when aggregate performance improves, reinforcing the need for evaluations that explicitly measure unintended consequences and failure patterns. (9) [9] Cognitive biases such as anchoring, where early information disproportionately shapes later judgment, are particularly salient for LLM-based tools because model outputs may provide a compelling “first narrative” that clinicians then refine rather than independently reconstruct. (10) [10]

Recent trial evidence underscores that simply providing clinicians access to an LLM does not guarantee better performance. In a randomized clinical trial evaluating LLM access during diagnostic reasoning, physicians given an LLM did not significantly outperform those using conventional resources, while the LLM alone achieved higher performance than physician groups, suggesting that collaboration failure can arise even when the model itself is strong. (11) [11] This gap between model capability and clinical benefit motivates the study of interaction protocols and workflow design. Human-centered design guidance for AI systems emphasizes setting expectations, supporting user correction, and designing for inevitable error and recovery over time, principles intended to foster appropriate reliance and reduce harm when systems are wrong. (12) [14]

Our prior work offers a concrete demonstration that workflow design can convert LLMs from passive tools into active collaborators. In a randomized trial of clinician–AI collaborative diagnostic workflows, structured collaboration protocols improved clinician accuracy compared with traditional resources, indicating that benefit can be unlocked by redesigning how clinicians and models exchange reasoning, rather than by model performance alone. (13) [12] Extending this interaction-centered lens, our subsequent work shows that clinician input can function as a powerful steering force for frontier models: exposure to clinician reasoning can increase concordance and improve some outcomes, while adversarial or incorrect clinician context can degrade outputs and surface vulnerabilities that are interaction-specific. (14) [13] Together, these findings motivate the central premise of the present study: for oncology evidence-support tools, the most informative evaluation must measure not only the quality of model outputs in isolation, but also the *clinical workflows* in which they are embedded, the *interaction patterns* clinicians adopt, and the *frictions* and *failure modes* shaping trust and use.

Accordingly, this study is designed as a mixed-methods evaluation centered on oncologist interaction with LLM-based evidence tools used in clinical practice. The quantitative component assesses clinician-rated utility of AI-generated literature review reports across multiple LLM-backed products, with prespecified outcomes and modeling approaches. The qualitative component uses semistructured interviews and free-response prompts to capture how oncologists currently use LLMs (including systems such as ChatGPT and OpenEvidence), which workflows they prioritize, where friction points occur (e.g., time, provenance, citation quality, integration into decision-making), and what improvements clinicians would recommend to enhance product usefulness and safety. This qualitative focus is formalized through a prespecified transcript parsing prompt that guides structured extraction of key themes, actionable recommendations, and participant-reported experiences.

A mixed-methods approach is well suited to studying LLM tools in oncology because utility is not purely an accuracy metric: it includes perceived credibility, time savings, cognitive burden reduction, workflow fit, and the degree to which outputs enable confident action. Mixed-methods reporting guidance emphasizes that integration across quantitative and qualitative components must be transparent to support interpretability and reproducibility. (15) [15] For the qualitative component, reporting standards such as COREQ highlight the importance of transparent methods describing context, data collection, and analytic procedures, including how themes are derived and supported by quotations. (16) [16] The study's qualitative analysis strategy is grounded in thematic analysis principles, which provide a systematic framework for developing themes from interview data while remaining flexible to the complexities of clinical experience. (17) [17]

In addition to evaluating LLMs as clinical tools, the present study advances a novel methodological contribution: using LLMs as structured analytic instruments for interview and transcript processing via prespecified prompts. Traditional qualitative analysis is time-intensive and can be influenced by analyst subjectivity, reflexivity, and variable coding decisions. LLM-assisted qualitative analysis has shown promise in healthcare-adjacent contexts, where older generation models can reproduce many high-level themes identified by human analysts. (18,19) [18] These findings suggest that LLMs can support qualitative workflows when used with clear guardrails and human oversight, but that rigorous methods are needed to ensure transparency and prevent "black box" theme generation.

Pre-specification is crucial for such an LLM analysis. In a large-scale evaluation of model-based sentiment judgments, some models approached human inter-rater reliability while others did not, and performance differences persisted even with tuning—demonstrating that model choice and analytic design materially affect measurement validity. (20) [19] Translating this insight to oncology interview research, prespecified prompts can be treated as standardized measurement protocols: they constrain analytic degrees of freedom, support auditability of outputs, and reduce the risk that qualitative findings reflect opportunistic prompt iteration or selective extraction. The prespecified transcript parsing prompt used in this study operationalizes these goals by instructing consistent extraction of practice patterns, friction points, and improvement opportunities from transcripts while excluding interviewer contributions and protecting confidentiality through anonymization directives.

Finally, the clinical translation of LLM-based evidence tools requires attention to ethics, governance, and regulation, particularly as these systems increasingly influence clinician cognition and decision flow. Global guidance emphasizes that AI in health should put ethics and human rights at the center of design and deployment, including accountability to healthcare workers who rely on these technologies. (21) [21] In the U.S., updated federal guidance on clinical decision support software clarifies regulatory scope and underscores the importance of transparency about the information presented to healthcare professionals. (22) [22] Within this landscape, a workflow-grounded, mixed-methods evaluation of oncology clinicians' real LLM use, and a transparent approach to LLM-assisted transcript measurement, can generate actionable evidence for improving model behavior, product design, and safe integration into clinical practice.

1.1 Study Aims

Aim 1 (primary): Estimate and test the difference in perceived overall utility between the custom OpenEvidence report and the standard OpenEvidence report across vignette evaluations, and other included report types.

Aim 2 (secondary/ mixed-methods):
- Quantitatively compare subcategory ratings across all four report types.
- Quantify stated "best" and "worst" report preferences and summarize preferred report length characteristics. (SAP_summary.docx)
- Qualitatively synthesize improvement requests, strengths/weaknesses, and cross-cutting tradeoffs using prespecified prompt-based pipelines on free responses and interview transcripts.

1.2 Study Hypotheses

1.2.1 Primary Hypotheses

H1 - The custom OpenEvidence report, ranking evidence levels, has higher mean overall utility rating than the standard OpenEvidence report, as measured on the 7-point Likert utility item and estimated from a mixed-effects model accounting for repeated measures within participants and vignettes.

1.2.2 Secondary Hypotheses

Secondary comparisons:

- Differences in mean utility between each other report type and standard OpenEvidence.
    - Medical specialist models will have greater percieved utility than generalist models
- Subcategory Likert ratings differ by report type for each
    - Medical specialist models (OpenEvidence, DoxGPT) will have greater scores relating to evidence synthesis. Generalist model (chatgpt) will have better readability
- Best/worst report choices are not uniform across report types (departure from 25% each)
    - Medical specialist models will be more favored than generalist models
- Optimal report length will be much shorter than given across all assessed reports

#### 2. Study Population

2.1 Inclusion Criteria
Participants are physicians with oncology expertise: licensed physicians with at least PGY2 completed, capacity to provide informed consent, access to necessary internet/technology for remote participation, and time availability for ~1 hour of tasks; there is a stated preference (not strict requirement) for specialty oncology training and specifically to fellows (e.g., hematology-oncology, radiation oncology).

2.2 Exclusion Criteria
Not fluent in English

2.3 Data Acquisition
Fill in all relevant information:

| |
|---|
| Study design: Remote, within-participant, randomized presentation order vignette evaluation ("randomized vignette study") with four randomized AI report types presented per vignette and blinded report source labeling within the survey instrument |
| Data source / how the data were collected: Qualtrics, interview transcripts |
| Contact information for team member responsible for data collection / acquisition: Bryan Bunning |
| Data or version (if downloaded, provide date): n/a |
| Data transfer method and date: n/a |
| Where dataset is stored: Qualtrics |

Notes:
Description:
Participants will see up to 5 oncology literature review vignettes in the survey; each vignette includes four blinded AI responses (OpenEvidence, Doximity, ChatGPT, and a custom prompt developed by the study team), with vignette order and AI response order randomized. Missing data will be a function of time, as the survey is hard capped at 1 hour for both the vignettes and qualitative interview.

#### 3. Outcomes, Exposures, and Additional Variables for Interest.

3.1 Primary Outcome(s)

| Outcome | Description | Variables and source | Specifications |
|---|---|---|---|
| Overall utility (7-point Likert) | Participant-rated overall utility of a vignette-specific AI report. Specifically our custom report vs OpenEvidence | Survey response in Qualtrics during session | Numeric coded 1–7 Measured once per report type per vignette, immediately after review of that report |

3.2 Secondary Outcome(s)

| Outcome | Description | Variables and source | Specifications |
|---|---|---|---|
| Utility contrasts vs OpenEvidence | Same utility item used for primary, additionally comparing other report types vs OpenEvidence | Same as primary | Derived from mixed model fixed effects for report type indicators. |
| Subcategory ratings (6 domains; each 7-point Likert) | Participant ratings on six subcategory items | Survey responses | Six separate Likert items; domain names and anchors unspecified; analyzed per domain. |
| Best report type | Participant selection of "best" report type | Survey response | Categorical with 4 levels (report types). collected |

| | | | |
|---|---|---|---|
| | | | overall at end of study. |
| Worst report type | Participant selection of "worst" report type | Survey response | Same structure as best report. |
| Optimal report length | Participant-stated preferred/optimal report length | Survey response | Distributional descriptive endpoint; page length. |

3.3 Additional Variables of Interest

| Outcome | Description | Variables and source | Specifications |
|---|---|---|---|
| n/a | | | eg, how is it coded? |

#### 4. Statistical Analysis Plan

4.1 Demographic and Clinical Characteristics ("Table 1")
Table 1 will describe the participant cohort using the variables actually collected.
Minimum Table 1: institution, training level category (resident / fellow / attending).
?Expanded Table 1: years of practice, oncology focus, current AI tool usage, and self-assessed AI risk tolerance.
Continuous variables (if present) will be summarized using SMD, categorical as count and percentages.

4.2 Analysis Plan for Aim 1
Aim 1 corresponds to the prespecified primary endpoint: overall utility.

Populations:

- Full analysis set (FAS): all consented participants who begin the Qualtrics instrument and provide at least one overall utility rating.
- Qualitative set: participants who complete an exit interview with audio recording consent (audio recording is optional per IRB).

Model specification (primary):
Primary model: linear mixed-effects model for overall utility rating with fixed effects for report type and random intercepts for participant and vignette, with standard OpenEvidence as the reference category: -

$Utility_{ijr} = \beta 0 + \beta 1 \cdot I(ChatGPT) + \beta 2 \cdot I(Doximity) + \beta 3 \cdot I(CustomOE) + u_{participant\_i} + u_{vignette\_j} + \varepsilon_{ijr}$
where $u_{participant\_i} \sim N(0, \sigma_p^2)$, $u_{vignette\_j} \sim N(0, \sigma_v^2)$, $\varepsilon_{ijr} \sim N(0, \sigma^2)$. REML utilized.

Justification note (methodological): Treating Likert as continuous is common for 7-point scales.

Sensitivity/augmentation: Add training level as a fixed effect

Aim 1 includes one prespecified primary hypothesis test (CustomOE vs OE). This SAP controls type I error at $\alpha=0.05$ for that single primary test.

*Missing data handling (Aim 1)*
Expected sources: incomplete vignette completion, skipped ratings, technical disruptions in remote

session, and optional refusal of audio recording (qualitative only).
Primary approach: mixed-effects modeling uses all available observed utility ratings under a Missing At Random (MAR) assumption conditional on model terms (participant/vignette random effects and included covariates).
Missingness description: report item-level missingness per report type and per vignette. Assumption that participants will not complete every vignette- randomization utilized to address potential bias.

4.2 Analysis Plan for Aim 2
Utility contrasts vs OpenEvidence (other report types):
From the Aim 1 mixed model, report β1 (ChatGPT vs OE) and other report β, including 95% CIs and multiplicity-adjusted p-values as specified below.
Subcategory Likert ratings:
The same structure of mixed effects model will be used per subcategory, with unadjusted p values

Best and worst report selections:

The SAP summary specifies reporting percentages and a chi-squared goodness-of-fit test against a 25% uniform choice model for best and worst separately. This SAP operationalizes:

- If best/worst is collected once per participant overall: one multinomial count vector (n participants) tested via chi-squared goodness-of-fit and supplemented with exact multinomial confidence intervals.

Optimal report length:
Summarize and visualize the distribution. Categorical: report proportions and plot bar chart.

*Sample size and assumptions*
Target enrollment: 30 clinicians with prior oncology experience (consent) and protocol expectation “up to 30.”

Protocol justification: A power calculation found n=23 sufficient to detect a difference between 50% and 75% preference toward the custom report compared with standard OpenEvidence; the protocol therefore targets up to 30 to allow for attrition and additional precision.

*4.3* Qualitative analysis plan

Data sources: free-text comments from the Qualtrics survey and exit interview transcripts (audio-recorded with permission).

Approach: Prompt-guided qualitative synthesis using prespecified workflows (SEE APPENDIX):
- Transcript-level inductive coding and anonymized segment extraction (transcript_parse_prompt).
- Cross-transcript synthesis into higher-order themes (transcript_synthesis).
- Sentiment feature extraction for pooled free responses with structured JSON outputs (sentiment_features).
- Second-order cross-report synthesis across JSON outputs (sentiment_cross_report). (

To better explain the LLM analysis:

- Every interview transcript will be put individually into transcript_parse_prompt to create a structured dataset of information gleaned from the interview
- All transcript structured datasets will be combined and fed into transcript_synthesis to yield study wide reccomendations
- All survey data (free response questions during the survey) will be combined **<u>*per report color*</u>** and fed into sentiment_features. This will yield 4 total structured datasets. Each dataset will include the specific advice for each colored report.
- All colored report datasets will be fed into sentiment_cross_report to yield the generalized advice originating from the survey data.

JSONs will be made publicly available

Qualitative recommendations for improvement in manuscript will be derived from transcript synthesis JSON after assessment from coauthors. Coauthors will have discretion in interpretation of summarized information.

Coauthors will suggest the features included in a next generation report style incorporating feedback derived from JSON data.

#### 5. Limitations

We are not assessing medical correctness! This is a massive limitation. The study downstream of correctness and is an HCI in conveying literature information to a physician. As a reminder, the majority of US doctors (Offcall survey, OE internal reporting) already using these tools!!
The framing of discussion is about AI literature review because these tools are not making medical decisions, they are assisting an Oncologist with a medical decision. We are not replacing oncologists with these tools.
Oncology specialty doesn't match to doctors
Bias to academic institutions

#### 6. Addendum for Additional Analyses

[enter if applicable]

#### 7. Appendix

TRANSCRIPT_PARSE_PROMPT

You will analyze ONE interview transcript from a mixed-methods study about AI-generated literature review report generation for oncology care workflows.

STUDY GOAL:

Identify themes, likes, dislikes, and actionable improvements for AI literature-review report generation that would increase utility, trust, and fit within oncology clinical workflows.

INDUCTIVE ANALYSIS REQUIREMENT:

Use an inductive approach: derive codes and themes from what the PARTICIPANT (oncologist) says in this transcript. Do NOT impose external frameworks (e.g., TAM/UTAUT) unless the participant explicitly invokes them. Do NOT use prior transcripts, general oncology knowledge, or assumptions.

INTERVIEWER EXCLUSION RULE (CRITICAL):

Bryan Bunning is the interviewer. His comments/questions are NOT data for thematic analysis.

- Exclude Bryan Bunning's statements from coding, themes, likes/dislikes, requirements, and memos.
- You MAY use interviewer prompts only to understand what the participant is responding to, but do not treat them as evidence.
- If speaker labels are missing, treat any question-like, leading, or explanatory text as "interviewer" and exclude it unless the participant repeats/endorses it in their own words.

ANONYMIZATION RULE (CRITICAL):

Preserve participant anonymity in the JSON output.

- Do NOT include any personal names of participants, colleagues, patients, family members, staff, or institutions/locations that could identify a person (e.g., specific hospitals/clinics, unique teams) anywhere in the JSON.
- Do NOT include participant initials, email addresses, phone numbers, usernames, exact addresses, or other identifiers.
- Replace any identifying proper nouns in participant excerpts with bracketed placeholders while keeping meaning:
  - Person → "[NAME]"
  - Patient → "[PATIENT]"
  - Hospital/Clinic/Institution → "[INSTITUTION]"
  - City/State/Country or other location → "[LOCATION]"
  - Unique program/unit/team → "[SERVICE]"
  - Email/phone/ID → "[IDENTIFIER]"
- Tool/product names that are not identifying individuals (e.g., Epic, OpenEvidence, NCCN, SecureGPT) may be kept unless the participant ties them to a uniquely identifying local instance; if so, replace with "[SYSTEM]".
- If a quote cannot be anonymized without losing its meaning, set representative_quote to null and explain briefly in the segment rationale that it was withheld for anonymization (do not add extra JSON keys).
- transcript_id must be anonymized (no names); use a non-identifying ID like "TX001" or a hash-like string you invent.

UNIT OF ANALYSIS:

A "meaningful segment" of participant speech (1–6 sentences) that expresses a single idea or closely related ideas. Use timestamps if available.

IMPORTANT CONSTRAINTS:

- Use ONLY information present in the transcript.

- Do NOT add external oncology knowledge or guidelines not explicitly mentioned by the participant.
- Do NOT invent quotes, speakers, motivations, or clinical facts.
- Quotes must be verbatim participant excerpts (no paraphrasing as quotes) EXCEPT for required anonymization placeholders as specified above.
- If a point is ambiguous, label it “uncertain/ambiguous” and explain why.
- If the transcript does not contain enough evidence for a category, output “Not observed in this transcript.”

INPUT TRANSCRIPT:
Transcript is attached
(If the transcript includes timestamps, preserve them.)
OUTPUT FORMAT (STRICT):
Return JSON ONLY, matching the schema below exactly (no extra keys, no commentary).
JSON SCHEMA:
{
"transcript_id": "[ANONYMIZED ID YOU PROVIDE]",
"interview_context": {
"participant_role": "oncologist|fellow|attending|unknown",
"setting_or_workflow_context": "free text or 'unknown' (must be non-identifying; avoid named places/institutions)",
"tools_mentioned_by_participant": ["list of tools/systems mentioned verbatim by participant, if any (non-identifying tools ok)"]
},
"segments": [
{
"segment_id": "S1",
"timestamp_start": "if available else null",
"timestamp_end": "if available else null",
"speaker": "participant",
"verbatim_excerpt": "exact participant text from transcript (<=60 words), anonymized with placeholders if needed",
"codes": ["code_1", "code_2"],
"valence": "positive|negative|mixed|neutral",
"object_of_evaluation": "e.g., length, tone, evidence display, tables, citations, workflow fit, trust, UI, visuals, guideline content, etc.",
"rationale": "1–2 sentences explaining why you assigned these codes using ONLY the excerpt (may note anonymization if quote withheld)."
}
],
"codebook": [
{
"code": "string_snake_case",
"definition": "operational definition grounded in participant language (1–2 sentences)",
"inclusion_criteria": ["bullets"],
"exclusion_criteria": ["bullets"],
"example_segment_ids": ["S1","S7"]
}
],

"themes": [
{
"theme_id": "T1",
"theme_name": "short descriptive name using participant-near language when possible",
"theme_definition": "2–4 sentences grounded in participant statements (no outside concepts)",
"supporting_segments": ["S1","S3","S9"],
"representative_quotes": [
{
"segment_id": "S1",
"quote": "verbatim participant quote (<=35 words), anonymized with placeholders if needed, or null if cannot be safely anonymized"
}
],
"overall_valence": "positive|negative|mixed|neutral",
"confidence": "high|medium|low",
"confidence_basis": "e.g., repeated across multiple participant segments vs single mention"
}
],
"likes": [
{
"like_id": "L1",
"statement": "concise statement of what the participant liked",
"supporting_segments": ["S2","S8"],
"representative_quote": "verbatim participant quote (<=25 words), anonymized with placeholders if needed, or null if cannot be safely anonymized"
}
],
"dislikes": [
{
"dislike_id": "D1",
"statement": "concise statement of what the participant disliked",
"supporting_segments": ["S4"],
"representative_quote": "verbatim participant quote (<=25 words), anonymized with placeholders if needed, or null if cannot be safely anonymized"
}
],
"requirements": [
{
"req_id": "R1",
"type": "must_have|nice_to_have|avoid",
"requirement": "single sentence requirement phrased as a product/design constraint derived from participant statements",
"supporting_segments": ["S4","S10"],
"risk_if_ignored": "1 sentence"
}
],
"workflow_integration": {
"current_workarounds": ["participant-described alternatives/workarounds, if any"],

"intended_use_cases": ["participant-described use cases, if any"],
"constraints": ["participant-mentioned constraints: time, cognitive load, trust, documentation, etc."],
"output_format_preferences": ["participant-mentioned formatting preferences"]
},
"trust_and_safety": {
"trust_drivers": ["participant-mentioned trust builders"],
"trust_erosions": ["participant-mentioned trust reducers"],
"verification_behaviors": ["participant-described checking behaviors, if mentioned"],
"medico_legal_or_policy_concerns": ["participant-mentioned concerns, else empty"]
},
"quantified_mentions": {
"top_codes_by_count": [
{"code": "string_snake_case", "count": 0}
],
"notes": "Counts are approximate; note any ambiguity due to transcript format and anonymization replacements."
},
"analytic_memos": {
"summary": "5–8 sentences summarizing the participant perspective only (must be non-identifying; no names/places).",
"tensions_or_tradeoffs": ["tradeoffs explicitly or implicitly described by the participant"],
"open_questions_for_next_interview": ["2–5 probes based on what this participant raised"]
}
}

CODING GUIDANCE:

- Only code PARTICIPANT speech.
- Use inductive, descriptive codes that stay close to participant wording.
- Create new codes when new ideas emerge; reuse existing codes within the transcript when the same idea recurs.
- Keep the codebook typically 10–25 codes per transcript.

Now produce the JSON.

Transcript_synthesis:
You will analyze a SET of JSON summaries generated from individual interview transcripts in a mixed-methods study about AI-generated literature review report generation for oncology care workflows.

**STUDY GOAL**

Synthesize findings across multiple oncologist interviews to identify cross-participant themes, recurring likes and dislikes, workflow needs, trust considerations, and design implications for AI-generated literature review reports used in oncology care.

**ANALYTIC TASK**

You are performing a **cross-case qualitative synthesis** using the structured JSON outputs from individual interviews as the sole input data source.
Your task is to:

- Compare patterns across participants
- Aggregate overlapping codes, likes, dislikes, requirements, workflow constraints, and trust/safety observations
- Identify higher-order synthesized themes grounded in the interview JSONs
- Produce an executive summary and publication-ready synthesized statements
- Preserve nuance, including disagreement, minority viewpoints, and tensions/tradeoffs

**INDUCTIVE SYNTHESIS REQUIREMENT**

Use an inductive approach. Build synthesis from the content of the provided interview JSONs only.

- Do NOT introduce external frameworks (e.g., TAM, UTAUT, human factors models) unless they are explicitly present in the interview outputs.
- Do NOT add outside oncology knowledge, product design assumptions, or clinical workflow assumptions not reflected in the input JSONs.
- Do NOT infer participant intent beyond what is reasonably supported by the source JSONs.
- Do NOT collapse distinct ideas into one theme if meaningful differences remain.

**INPUT DATA RULES**

The input consists of multiple interview-summary JSON objects, each derived from one participant interview.
Assume each JSON may contain:

- segments
- codebook
- themes
- likes
- dislikes
- requirements
- workflow_integration
- trust_and_safety
- quantified_mentions
- analytic_memos

Use these interview JSONs as the analytic record.

**INTERVIEWER EXCLUSION RULE**

Bryan Bunning is the interviewer and is not part of the analyzed participant data.
If any interviewer-originated ideas appear to have been improperly carried into an interview JSON, do not elevate them unless clearly endorsed by participant-derived evidence in that JSON.

**UNIT OF SYNTHESIS**

The primary unit of synthesis is the **participant-level interview JSON**.
The secondary units are:

- coded segments

- interview-level themes
- likes/dislikes
- requirements
- workflow/trust observations

**IMPORTANT CONSTRAINTS**

- Use ONLY information present in the provided interview JSONs.
- Do NOT invent quotations.
- Do NOT create false consensus.
- When participants differ, explicitly describe the divergence.
- Preserve minority but salient viewpoints if they have clear implications.
- Distinguish:
    - commonly mentioned findings
    - strongly expressed findings
    - design-critical findings
- If support is weak or inconsistent, label it accordingly.
- If a category is not sufficiently supported across interviews, output: "Insufficient cross-interview evidence."

**SYNTHESIS GUIDANCE**

When synthesizing:

- Merge semantically overlapping codes into broader synthesized concepts only when justified by the source material.
- Preserve original participant-near language when possible.
- Note the number of interviews contributing to a theme.
- Treat frequency as informative but not definitive; a less frequent issue may still be important if it is concrete, high-risk, or strongly stated.
- Pay special attention to:
    - workflow fit
    - time burden
    - trust and verification
    - evidence display and citations
    - report organization
    - visual formatting
    - clinical usefulness
    - cognitive load
    - ambiguity tolerance
    - medico-legal concerns
    - perceived risks of misleading output
    - desired safeguards and product requirements

**INPUT**

[PASTE ARRAY OF INDIVIDUAL INTERVIEW JSON OBJECTS HERE]

**OUTPUT FORMAT**

Return JSON ONLY, matching the schema below exactly.

{
"synthesis_metadata": {
"study_focus": "AI-generated literature review report generation for oncology care workflows",
"num_interviews_synthesized": 0,
"num_interviews_with_usable_data": 0,
"notes_on_input_quality": "Brief note on missingness, inconsistent detail, or other limitations across

interview JSONs."
},
"executive_summary": {
"summary_paragraph": "6-10 sentence publication-ready summary synthesizing the overall findings across interviews.",
"top_line_findings": [
"3-7 concise high-level findings suitable for a results section or abstract"
],
"overall_interpretation": "2-4 sentences interpreting what these findings suggest about the design and implementation of AI literature review tools in oncology workflows, grounded only in the input data."
},
"cross_interview_themes": [
{
"synth_theme_id": "ST1",
"theme_name": "short descriptive name",
"theme_definition": "3-5 sentences describing the synthesized theme across interviews",
"contributing_interviews": ["transcript_id_1", "transcript_id_2"],
"num_interviews_supporting": 0,
"supporting_evidence": {
"supporting_theme_ids": ["T1", "T2"],
"supporting_segment_ids": ["transcript1:S3", "transcript4:S8"],
"supporting_like_ids": ["transcript2:L1"],
"supporting_dislike_ids": ["transcript3:D2"],
"supporting_requirement_ids": ["transcript5:R1"]
},
"representative_quotes": [
{
"transcript_id": "string",
"segment_id": "S1",
"quote": "verbatim quote <=35 words"
}
],
"direction_of_sentiment": "positive|negative|mixed|neutral",
"prevalence": "high|moderate|low",
"importance": "high|moderate|low",
"rationale_for_importance": "Explain whether this theme matters because of recurrence, strength of language, workflow consequences, trust implications, or design risk.",
"variation_or_disagreement": "Describe important differences across participants, or state 'Minimal disagreement observed.'",
"confidence": "high|medium|low",
"confidence_basis": "Explain confidence based on consistency, richness, and number of supporting interviews."
}
],
"synthesized_likes": [
{
"s_like_id": "SL1",
"statement": "Cross-interview synthesized statement of what participants valued",

"contributing_interviews": ["transcript_id_1", "transcript_id_3"],
"num_interviews_supporting": 0,
"example_quotes": [
{
"transcript_id": "string",
"segment_id": "S2",
"quote": "verbatim quote <=25 words"
}
]
}
],
"synthesized_dislikes": [
{
"s_dislike_id": "SD1",
"statement": "Cross-interview synthesized statement of what participants disliked",
"contributing_interviews": ["transcript_id_2", "transcript_id_4"],
"num_interviews_supporting": 0,
"example_quotes": [
{
"transcript_id": "string",
"segment_id": "S5",
"quote": "verbatim quote <=25 words"
}
]
}
],
"design_requirements": [
{
"design_req_id": "DR1",
"priority": "must_have|should_have|nice_to_have|avoid",
"requirement": "Single sentence product/design requirement synthesized across interviews",
"why_it_matters": "1-2 sentences grounded in participant evidence",
"contributing_interviews": ["transcript_id_1", "transcript_id_2"],
"num_interviews_supporting": 0,
"risk_if_ignored": "1 sentence",
"supporting_evidence": ["transcript1:R2", "transcript3:D1", "transcript4:S7"]
}
],
"workflow_synthesis": {
"intended_use_cases": [
{
"use_case": "synthesized use case",
"contributing_interviews": ["transcript_id_1"],
"num_interviews_supporting": 0
}
],
"workflow_constraints": [
{

"constraint": "synthesized workflow constraint",
"contributing_interviews": ["transcript_id_2"],
"num_interviews_supporting": 0
}
],
"format_preferences": [
{
"preference": "synthesized output-format preference",
"contributing_interviews": ["transcript_id_3"],
"num_interviews_supporting": 0
}
],
"current_workarounds": [
{
"workaround": "participant-described workaround synthesized across interviews",
"contributing_interviews": ["transcript_id_4"],
"num_interviews_supporting": 0
}
]
},
"trust_and_safety_synthesis": {
"trust_drivers": [
{
"driver": "synthesized trust builder",
"contributing_interviews": ["transcript_id_1", "transcript_id_5"],
"num_interviews_supporting": 0
}
],
"trust_erosions": [
{
"erosion": "synthesized trust reducer",
"contributing_interviews": ["transcript_id_2"],
"num_interviews_supporting": 0
}
],
"verification_behaviors": [
{
"behavior": "how participants described checking or validating outputs",
"contributing_interviews": ["transcript_id_3"],
"num_interviews_supporting": 0
}
],
"medico_legal_or_policy_concerns": [
{
"concern": "synthesized concern",
"contributing_interviews": ["transcript_id_4"],
"num_interviews_supporting": 0
}

]
},
"tensions_and_tradeoffs": [
{
"tradeoff_id": "TT1",
"tradeoff": "Concise statement of a tension observed across interviews",
"side_a": "first side of the tension",
"side_b": "second side of the tension",
"contributing_interviews": ["transcript_id_1", "transcript_id_2"],
"interpretation": "1-3 sentences explaining the tension in participant-grounded terms"
}
],
"publication_ready_synthesized_statements": [
{
"statement_id": "P1",
"type": "results|discussion|implication",
"statement": "A polished, publication-ready synthesized statement grounded in the data and suitable for a manuscript.",
"support_level": "strong|moderate|preliminary",
"basis": "Brief explanation of support based on breadth/consistency/richness of interview evidence."
}
],
"minority_or_divergent_views": [
{
"view_id": "MV1",
"statement": "Important but less common or divergent viewpoint",
"contributing_interviews": ["transcript_id_6"],
"why_it_should_be_retained": "Why this viewpoint remains analytically important despite lower prevalence"
}
],
"evidence_map": {
"most_salient_synthesized_codes_or_concepts": [
{
"concept": "string",
"approx_num_interviews": 0,
"related_interview_codes": ["code_a", "code_b"]
}
],
"notes": "Counts are approximate and based on interview-level JSON outputs rather than re-coding full transcripts."
},
"methodological_memo": {
"cross_case_summary": "5-8 sentences summarizing the analytic pattern across participants.",
"areas_of_strong_convergence": [
"list"
],
"areas_of_partial_convergence": [

"list"
],
"areas_of_disagreement_or_heterogeneity": [
"list"
],
"limitations_of_this_synthesis": [
"Important limitations caused by input quality, missing detail, uneven coding depth, or ambiguity."
]
}
}

Sentiment_features:
You will analyze a concatenated set of qualitative comments from practicing oncologists about AI-generated reports and literature-review outputs used in oncology workflows.

**STUDY PURPOSE**

The purpose of this analysis is to identify the **strengths, weaknesses, and improvement opportunities** described by the cohort regarding AI-generated reports and literature-review tools for oncology practice.

**UNIT OF ANALYSIS**

The input consists of **multiple short free-text comments concatenated together** from one analytic subset, such as:

- one **report type** within a vignette,
- one **report type across multiple vignettes**, or
- **study closeout comments** aggregated across participants.

Treat the full input as a **pooled corpus of comments from multiple oncologists**, not as a single coherent narrative from one speaker.

**ANALYTIC TASK**

Extract and synthesize the **positive features**, **negative features**, and **suggested improvements** mentioned in the text.
Your goal is to produce a structured summary of:

1. what oncologists liked,
2. what oncologists disliked,
3. what changes they wanted,
4. and which issues appeared most salient in this subset.

**INDUCTIVE ANALYSIS REQUIREMENT**

Use an **inductive qualitative approach**:

- Derive categories from the language and content of the comments themselves.
- Do **not** impose external frameworks unless explicitly supported by the text.
- Do **not** infer attitudes that are not grounded in the input.
- Stay close to the wording and meaning used by the participants.

**IMPORTANT INTERPRETIVE RULES**

- These comments may be fragmented, telegraphic, redundant, or inconsistently phrased.
- Normalize minor spelling/grammar issues mentally, but preserve the intended meaning.
- Multiple comments may express the same idea using different wording; consolidate these into a shared feature when appropriate.
- Distinguish between:
  - **positive features** (what worked well),
  - **negative features** (what did not work well),
  - **improvement requests** (what should be changed),
  - and **mixed/conditional sentiments** (e.g., “good content but too long”).
- When a comment contains both praise and criticism, capture both separately.
- Do not over-aggregate distinct concepts into one broad bucket if the text supports finer distinctions.

**REPORT-TYPE AWARENESS**

The input belongs to a specific analytic subset defined by the researcher, usually a **report type**.
You should summarize sentiment **within this subset only** and not compare to other report types unless the input explicitly includes such comparisons.
If the input is from study closeout questions rather than one report type, apply the same logic and summarize the pooled sentiment across the closeout corpus.

**OUTPUT REQUIREMENTS**

Return valid JSON only.

Use the following schema exactly:

```
{
 "analysis_unit": {
  "subset_type": "report_type | closeout | other",
  "subset_label": "string",
  "notes": "brief description of what this corpus represents"
 },
 "summary": {
  "overall_impression": "2-4 sentence synthesis of the major strengths and weaknesses in this subset",
  "dominant_positive_themes": ["theme 1", "theme 2", "theme 3"],
  "dominant_negative_themes": ["theme 1", "theme 2", "theme 3"],
  "dominant_improvement_requests": ["request 1", "request 2", "request 3"]
 },
 "positive_features": [
  {
   "feature_label": "concise name of positive feature",
   "description": "brief explanation grounded in the comments",
   "representative_examples": [
    "short paraphrase or brief quote fragment 1",
    "short paraphrase or brief quote fragment 2"
   ],
   "relative_salience": "high | medium | low"
  }
 ],
 "negative_features": [
  {
   "feature_label": "concise name of negative feature",
   "description": "brief explanation grounded in the comments",
   "representative_examples": [
    "short paraphrase or brief quote fragment 1",
    "short paraphrase or brief quote fragment 2"
   ],
   "relative_salience": "high | medium | low"
  }
 ],
 "improvement_requests": [
  {
   "request_label": "concise name of requested improvement",
   "description": "brief explanation of what participants wanted changed",
   "representative_examples": [
    "short paraphrase or brief quote fragment 1",
    "short paraphrase or brief quote fragment 2"
   ],
   "targeted_problem": "what weakness this request appears to address",
   "relative_salience": "high | medium | low"
  }
```

```
 ],
 "mixed_or_tradeoff_comments": [
  {
   "theme_label": "concise name of tension or tradeoff",
   "description": "brief explanation of the mixed sentiment",
   "representative_examples": [
    "short paraphrase or brief quote fragment 1"
   ]
  }
 ],
 "analytic_memos": {
  "interpretive_notes": [
   "brief memo 1",
   "brief memo 2"
  ],
  "data_quality_notes": [
   "note on fragmentation/redundancy/ambiguity if relevant"
  ]
 }
}
```

**CODING GUIDELINES**

When grouping comments, favor categories such as:

- clarity
- organization
- brevity vs verbosity
- explicit reasoning
- usefulness of citations
- citation specificity
- transparency of evidence use
- actionability for clinical use
- completeness
- trustworthiness
- workflow fit
- readability
- formatting/structure

Only use these categories if they are supported by the text. You may create other categories when the comments indicate a different concept.

**QUOTATION / EXCERPT RULES**

- Use only short excerpts or paraphrased fragments in representative_examples.
- Do not fabricate quotes.
- If wording is too fragmentary for a quote, paraphrase faithfully.
- Keep examples concise.

**SALIENCE RULES**

Estimate relative_salience based on:

- repetition across comments,
- clarity/directness of the sentiment,
- and emphasis in the language.

Do not treat salience as a numeric frequency count unless the data clearly supports that.

**DO NOT**

- Do not invent participant intent.
- Do not claim statistical prevalence.
- Do not compare subsets unless explicit in the input.
- Do not introduce external oncology knowledge.
- Do not overinterpret isolated fragments.
- Do not output anything except valid JSON.

Sentiment_cross_report

You will analyze **multiple JSON outputs** generated by the sentiment_features.docs prompt from a mixed-methods study of practicing oncologists evaluating AI-generated reports and literature-review outputs for oncology workflows.

**STUDY PURPOSE**

The purpose of this analysis is to synthesize, across report types or study sections, the major **strengths**, **weaknesses**, **requested improvements**, and **cross-cutting tradeoffs** that oncologists described regarding AI-generated reports and literature-review tools.

**INPUT**

The input consists of multiple structured JSON objects.

Each JSON object summarizes one analytic subset, such as:

- one **report type** within a vignette,
- one **report type across multiple vignettes**, or
- one **aggregated study closeout corpus**.

Each JSON contains:

- an analysis_unit,
- a synthesized summary,
- positive_features,
- negative_features,
- improvement_requests,
- mixed_or_tradeoff_comments,
- and analytic_memos.

**UNIT OF ANALYSIS**

Your job is to synthesize **across these subset-level summaries**.

This is a **second-order qualitative synthesis**:

- You are synthesizing across previously extracted themes/features.
- Do **not** return to line-by-line coding of raw participant comments.
- Work only from the provided JSON inputs.

**ANALYTIC TASK**

Produce a structured cross-report synthesis that identifies:

1. the most recurring positive features across subsets,
2. the most recurring negative features across subsets,
3. the most recurring improvement requests,
4. the most important tradeoffs/tensions,
5. points that appear common across report types,
6. and points that appear more subset-specific.

**INDUCTIVE SYNTHESIS REQUIREMENT**

Use an **inductive, evidence-grounded synthesis approach**:

- Build higher-order categories from the JSON inputs.
- Preserve distinctions when different subsets highlight meaningfully different issues.
- Consolidate overlapping labels when they clearly refer to the same underlying concept.
- Stay close to the content of the source JSONs.
- Do not introduce external frameworks unless explicitly justified by the input content.

**IMPORTANT INTERPRETIVE RULES**

- Treat salience as **qualitative recurrence and emphasis**, not as formal quantitative frequency.
- Do not make statistical claims such as “most oncologists” unless explicitly supported by the input.
- Look for both:

    - **cross-cutting themes** that recur across multiple subsets, and
    - **subset-specific themes** that appear more localized.
- When a theme appears in both positive and negative form across subsets, capture the tension explicitly.
    - Example: “comprehensive” may be valued by some comments but criticized as “too long” in others.
- Preserve nuance: do not flatten all themes into generic buckets if the subset-level JSONs contain clinically meaningful distinctions.

**OUTPUT REQUIREMENTS**

Return valid JSON only.

Use the following schema exactly:

```
{
 "corpus_summary": {
  "number_of_input_jsons": 0,
  "subset_labels": ["string"],
  "overall_synthesis": "4-6 sentence publication-ready synthesis describing the major strengths, weaknesses, and design implications across report types or study sections"
 },
 "cross_cutting_positive_features": [
  {
   "theme_label": "concise name of cross-cutting strength",
   "description": "brief synthesis of how this strength appeared across subsets",
   "appears_in_subsets": ["subset label 1", "subset label 2"],
   "qualitative_salience": "high | medium | low",
   "notes_on_variation": "optional note on how this strength differed by subset"
  }
 ],
 "cross_cutting_negative_features": [
  {
   "theme_label": "concise name of cross-cutting weakness",
   "description": "brief synthesis of how this weakness appeared across subsets",
   "appears_in_subsets": ["subset label 1", "subset label 2"],
   "qualitative_salience": "high | medium | low",
   "notes_on_variation": "optional note on how this weakness differed by subset"
  }
 ],
 "cross_cutting_improvement_requests": [
  {
   "theme_label": "concise name of recurring requested improvement",
   "description": "brief synthesis of what participants wanted changed across subsets",
   "appears_in_subsets": ["subset label 1", "subset label 2"],
   "targeted_problem": "what broader weakness this request seems intended to solve",
   "qualitative_salience": "high | medium | low"
  }
 ],
 "key_tradeoffs": [
  {
   "tradeoff_label": "concise name of recurring tension",
```

```
      "description": "brief explanation of the tradeoff across subsets",
      "positive_pole": "what users valued",
      "negative_pole": "what users found problematic",
      "appears_in_subsets": ["subset label 1", "subset label 2"]
    }
  ],
  "subset_specific_findings": [
    {
      "subset_label": "string",
      "distinctive_positive_features": ["feature 1", "feature 2"],
      "distinctive_negative_features": ["feature 1", "feature 2"],
      "distinctive_improvement_requests": ["request 1", "request 2"],
      "interpretive_note": "what seemed distinctive about this subset relative to the others"
    }
  ],
  "publication_ready_synthesis": {
    "results_paragraph": "1 paragraph suitable for a mixed-methods Results section describing cross-report feature patterns",
    "design_implications_paragraph": "1 paragraph suitable for Discussion or Implications describing what the findings suggest AI report tools should improve"
  },
  "analytic_memos": {
    "theme_merging_decisions": [
      "brief note explaining important theme consolidations"
    ],
    "uncertainties_or_limitations": [
      "brief note about ambiguity, inconsistent labeling, or limited comparability across subsets"
    ]
  }
}
```

**SYNTHESIS GUIDELINES**

When merging themes across subset-level JSONs:

- Merge labels that clearly reflect the same idea even if worded differently.
    - Example: “too long,” “verbose,” and “overly detailed” may be merged into a broader theme such as excessive length/verbosity.
- Keep distinct themes separate when they represent different concepts.
    - Example: lack of citation specificity should remain distinct from lack of overall trustworthiness unless the input clearly combines them.
- Favor labels that are:
    - concise,
    - publication-friendly,
    - and faithful to participant meaning.

**PRIORITY DOMAINS TO WATCH FOR**

Only use these when supported by the input:

- organization / structure
- length / concision
- explicit reasoning
- transparency

- citation specificity
- evidence traceability
- readability
- actionability
- completeness
- trustworthiness
- workflow fit
- interpretability
- clinical usefulness

**RESULTS PARAGRAPH STYLE**

The results_paragraph should read like a manuscript Results section:

- neutral and concise,
- theme-focused,
- no exaggerated claims,
- no unsupported quantification,
- no bullet formatting,
- appropriate for publication in a mixed-methods study.

**DESIGN IMPLICATIONS PARAGRAPH STYLE**

The design_implications_paragraph should translate the findings into practical implications for AI report design. It should focus on features that would likely improve utility for oncology workflows, based only on the input JSONs.

**DO NOT**

- Do not invent findings absent from the subset JSONs.
- Do not return to raw-data style quoting unless brief examples are already embedded in the source JSON logic.
- Do not make numerical prevalence claims unless explicitly supported by the inputs.
- Do not compare subsets beyond what the JSON evidence supports.
- Do not output anything except valid JSON.

ADDITIONAL PUBLICATION ORIENTATION:

- Prefer theme labels that would be understandable in a manuscript table or Results section.
- Emphasize recurring design priorities that cut across report types.
- Surface tensions that are especially relevant to clinical AI adoption, such as comprehensiveness versus brevity, or evidence synthesis versus citation traceability.
- Keep the tone analytical and restrained.
